# Effect of higher dose primaquine for the radical cure of *Plasmodium vivax* malaria in Indonesia: a systematic review and individual patient data meta-analysis

**DOI:** 10.1101/2025.11.13.25340059

**Authors:** Ihsan Fadilah, James A Watson, Ayodhia P Pasaribu, Inge Sutanto, Erni J Nelwan, Kartini Lidia, Megha Rajasekhar, Iqbal RF Elyazar, Walter RJ Taylor, Kamala Thriemer, Nicholas PJ Day, Jeanne Rini Poespoprodjo, Julie A Simpson, Ric N Price, J Kevin Baird, Robert J Commons

## Abstract

**Background:** *Plasmodium vivax* malaria has diverse transmission and relapse patterns in Indonesia. The optimal dose of primaquine to prevent relapses across the country is unknown. We evaluated the anti-relapse efficacy, gastrointestinal tolerability, and haematological safety (G6PD activity ≥30%) of different primaquine regimens in varied endemic settings in Indonesia.

**Methods:** We systematically searched for studies published between 1 January 2000 and 23 July 2024 prospectively enrolling patients with acute uncomplicated *P. vivax* malaria where some patients were treated with primaquine. Individual patient data (IPD) from eligible studies were pooled and harmonised. We fitted one-stage IPD multivariable regression models to estimate the causal relationship between the body weight-adjusted primaquine dose with three separate primary outcomes: (i) the time to first *P. vivax* recurrence (days 7–180), (ii) any gastrointestinal discomfort (days 5–7), and (iii) ≥25% reduction relative to baseline haemoglobin and a reduction to <7 g/dL (days 1–14).

**Findings:** Of ten eligible studies, seven were available for inclusion. Compared with a total dose of 3·5 mg/kg primaquine, patients treated with a total dose of 7 mg/kg had a lower rate of recurrence over 6 months (adjusted hazard ratio 0·53; 95% confidence interval [CI] 0·45 to 0·63; n = 1797); the relative efficacy was consistent across regions, but the absolute benefit varied. Gastrointestinal discomfort was more frequent with higher doses (adjusted risk ratio 1·32 per 0·25 mg/kg daily dose; 95% CI 1·15 to 1·51; n = 952). In 822 patients available to be assessed for haematological safety (788 [96%] with G6PD activity ≥70% and 34 [4%] with activity 30% to <70%), only one patient developed clinically relevant haemolysis.

**Interpretation:** Across all transmission settings in Indonesia, a total dose of 7 mg/kg halved the rate of recurrent *P. vivax* malaria over a 6-month period compared with the low dose of 3·5 mg/kg. However, increased daily doses slightly increased risks of gastrointestinal discomfort and haemolysis.

**Funding:** NDM Tropical Network Fund, Bill and Melinda Gates Foundation

**Research in context:** *Evidence before this study:* Indonesia is a geographically vast country with heterogeneous patterns of periodicity and varying magnitudes of risk for *Plasmodium vivax* relapse. However, the optimal regimen for the radical cure of *P. vivax* is unknown. The national antimalarial guidelines continue to recommend a low-dose regimen of 3·5 mg/kg without routine glucose-6-phosphate dehydrogenase (G6PD) testing. We systematically searched for trials conducted in Indonesia that randomly assigned patients with *P. vivax* malaria to primaquine dosing regimens. Articles published in any language between 1 January 2000 and 23 July 2024, using the terms "vivax" and "primaquine" in MEDLINE, Embase, Web of Science, Scopus, and the Cochrane Library were identified. No published randomised controlled trial directly comparing high-dose (≥5 mg/kg total) versus low-dose (2 to <5 mg/kg total) primaquine in Indonesia was identified. We identified two trials that randomised patients with *P. vivax* malaria to receive either high-dose (target total dose of 7 mg/kg over seven or 14 days) or low-dose (3·5 mg/kg over 14 days) primaquine, compared with placebo, in combination with dihydroartemisinin-piperaquine. One multi-country trial included two study sites in Sumatra: Hanura and Tanjung Leidong. Compared with placebo, high-dose primaquine reduced relapse by 60–89%. In another trial, patients acquired infections in Papua and were followed in malaria-free East Java. Low-dose primaquine reduced relapse by 74% compared with placebo.

*Added value of this study:* This country-specific systematic review and individual patient data meta-analysis included 1797 patients from seven studies evaluating anti-relapse efficacy, gastrointestinal tolerability, and haematological safety. The analysis represents the most comprehensive analysis to date of different primaquine dosing regimens for *P. vivax* malaria in Indonesia. The results suggest that doubling the total dose from 3·5 mg/kg to 7 mg/kg would halve the rate of *P. vivax* recurrence within 180 days, however, the absolute benefit of this reduction depends on the underlying risk of recurrence in different regions of Indonesia. An increase in the daily dose of primaquine resulted in a moderate increase in gastrointestinal discomfort; and only a rare occurrence of clinically relevant haemolysis, given that G6PD activity among patients was mostly ≥70%.

*Implication of all the available evidence:* For Indonesian patients with *P. vivax* malaria, a high total primaquine dose of 7 mg/kg is more efficacious than the currently implemented 3·5 mg/kg regimen. Higher daily doses of primaquine are associated with increased gastrointestinal discomfort, however, these may be mitigated by co-administration with food. Clinically relevant haemolysis is rare in patients with G6PD activity >30%.

## Background

*Plasmodium vivax* is more geographically widespread than other causes of human malaria. In regions where *P. vivax* and *P. falciparum* are co-endemic, *P. vivax* is increasingly the predominant species because conventional malaria control efforts more readily impact *P. falciparum*.^1^ As of 2024, nearly 80% of districts in Indonesia had achieved malaria-free status.^2^ However, *P. vivax* malaria remains prevalent with over 100,000 reported cases in recent years; most of them from the eastern Papua region.^3,4^

Dormant liver stages (hypnozoites) of *P. vivax* cause repeated blood-stage infections (relapses) and attacks of acute malaria that constitute a major cause of morbidity and mortality, particularly among young children and pregnant women.^5^ Relapsing *P. vivax* avoids conventional control efforts aimed at treating acute malaria and minimising mosquito vector contact. Radical cure of acute malaria, by combining blood schizontocidal drugs with an 8-aminoquinoline (primaquine or tafenoquine) that kills dormant hypnozoites, remains the only effective strategy to avert *P. vivax* relapses.

A meta-analysis of malaria studies across *P. vivax*-endemic regions globally published in 2024 suggested that a high total dose of 7 mg/kg can halve the rate of *P. vivax* recurrences compared with a low total dose of 3·5 mg/kg, with only a slight increase in gastrointestinal discomfort.^6^ Patients treated with a daily primaquine dose of up to 0·5 mg/kg with glucose-6-phosphate dehydrogenase (G6PD) activity >30% had comparable risks of haemolysis with patients not exposed to primaquine.^7^

Updated 2024 WHO guidelines reflected these findings, with a high total dose of primaquine recommended over either 14 days or 7 days.^8^ Indonesian national guidelines currently recommend a low-dose regimen of 3·5 mg/kg total over 14 days without routine G6PD testing, with a higher dose (7 mg/kg over 14 days) recommended for cases of suspected relapse or treatment failure.^9^ This conservative dosing strategy reflects ongoing operational challenges in implementing universal G6PD testing at scale within programme settings.^10^ However, Indonesia is a geographically large country with varied *P. vivax* epidemiological characteristics. For example, in Papua, Indonesia, there is a high rate of *P. vivax* transmission and high risk of relapse, even when high total dose primaquine is given,^11–13^ while other regions of Indonesia have reported good efficacy rates with low dose primaquine.^14,15^ Thus, the potential heterogeneity in effects, as well as the overall risks and benefits of different primaquine regimens for Indonesia, remains unclear. Currently, there is limited evidence from individual-level, head-to-head comparisons of primaquine regimens across regions in the country.

This study aimed to assess the anti-relapse efficacy, gastrointestinal tolerability, and haematological safety of the different primaquine doses in patients with *P. vivax* malaria in Indonesia.

## Methods

### Search strategy and selection criteria

Building on an existing living systematic review^16^ of studies of patients with acute uncomplicated *P. vivax* malaria, we conducted an updated systematic search of MEDLINE, Embase, Web of Science, Scopus, and the Cochrane Library for studies conducted in Indonesia and published in any language between 1 January 2000 and 23 July 2024. Search terms are provided in Supplementary Material (**List S*1***). Two reviewers (IF and RJC) conducted the review and resolved discrepancies through discussion, with JAW involved in cases of disagreement. The protocol was pre-registered with PROSPERO (CRD42024580630). We included randomised or non-randomised therapeutic trials with ≥28 days of active follow up for gastrointestinal tolerability and haematological safety, or ≥42 days for anti-relapse efficacy. Eligible studies included at least one group receiving multi-day primaquine for relapse prevention, initiated within seven days of blood schizontocidal treatment (chloroquine, quinine, or common artemisinin-based combination therapies).

### Data pooling

Investigators of eligible studies were invited to contribute individual patient data (IPD), including unpublished data when possible. Shared data were uploaded to the Worldwide Antimalarial Resistance Network repository and curated in accordance with the IDDO SDTM Implementation Guide.^17^ Patients were excluded if key variables were missing (age, sex, weight, baseline parasitaemia, treatment regimen), they had severe malaria, pregnancy, mixed infections, or received additional antimalarials after schizonticidal treatment. Additional inclusion criteria are described in Supplementary Material (**List S*2***). Included data were pseudo-anonymised and had received prior ethical approval locally. No additional approval was required, as per Oxford Tropical Research Ethics Committee guidance. This study is reported according to PRISMA-IPD guidelines^18^ (**Table S*1***).

### Endpoints

Day zero was defined as initiation of antimalarial treatment after enrolment. The primary efficacy endpoint was the time to the first *P. vivax* recurrence (ascertained by microscopy), irrespective of symptoms, between days 7–180. Secondary endpoints included the time to the first symptomatic recurrence and the number of recurrences within the same timeframe. The primary gastrointestinal tolerability endpoint was a composite endpoint including any of the following signs of gastrointestinal discomfort, ascertained by symptom questionnaire, occurring between days 5–7 (after the resolution of acute malaria symptoms and completion of schizonticidal treatment): vomiting, anorexia, diarrhoea. Secondary tolerability endpoints included similar symptoms on day 0 and days 1–2, and acute vomiting within 1 hour of primaquine administration. The primary haematological safety endpoint was a ≥25% haemoglobin reduction from baseline to <7 g/dL between days 1–14 (HemoCue™ system). Secondary endpoints included (i) maximum haemoglobin drop between days 1–14, 2–3, and 5–7, (ii) incident anaemia (Hb <11 g/dL) during days 2–3 in patients with baseline Hb ≥11 g/dL and normal G6PD activity (≥70% activity^19^; study subgroup of interest), and (iii) severe haemolytic adverse events (Hb <5 g/dL, >5 g/dL drop, blood transfusion, renal failure requiring dialysis, or death) within 14 days.

### Exposures

The efficacy exposure was the weight-adjusted (i.e., mg of primaquine base per kg of body weight [mg/kg]) total primaquine dose. The gastrointestinal tolerability and haematological safety exposure was the weight-adjusted daily dose.^6,7^ Doses were calculated from daily tablets or the mg administered and if these were unavailable they were inferred from protocols. Primaquine dose was specified in our models as a continuous variable. The effects of different categories of primaquine dose (**List S*2***) were explored.

### Data analysis

The main analyses estimated the effect of primaquine dose on recurrence, gastrointestinal tolerability, and haemolysis under a one-stage IPD meta-analysis framework.

#### Anti-relapse efficacy

Cumulative incidence of first recurrence was modelled with a multivariable Cox proportional hazards model including total primaquine dose, age, sex, natural log-transformed baseline parasite density, and study site. Study site (n = 6; Timika, Tanjung Leidong, Lumajang, Kupang, Sragen, Hanura) was specified as a stratification variable to allow each site to have its own baseline hazard function. A causal directed acyclic graph (DAG) guided model development (**Figure S*1***), and a restricted cubic spline for total primaquine dose allowed for nonlinearity (See **List S*2*** for further methodological details). We evaluated potential differential effects of total primaquine dose by age group (<5 years or ≥5 years) and origin of infection (within or outside Papua; reflecting a higher potential burden of hypnozoites and/or more primaquine-tolerant strains within Papua). Additional analyses, including sensitivity analyses and assessment of residual confounding using negative controls are described in Supplementary Material (**List S*2***).

#### Gastrointestinal tolerability

We summarised the outcome prevalence descriptively across study sites to highlight heterogeneity in the measurement of this endpoint. In the randomised subset of these data, the risk of gastrointestinal discomfort was modelled using multivariable Poisson regression^20^ with daily primaquine dose, age, sex, and log baseline parasite density (based on a DAG; **Figure S*3***), with dose categories and splines used to assess nonlinear effects. Study site was not included in the model, as the study population for this outcome originated from a single trial conducted under the same protocol. Similar model specification was used to model the risk of acute vomiting. As the outcome prevalence was high, Poisson regression was used to model the binary outcomes to estimate the risk ratio. Effect estimates from modified Poisson models^20^ were obtained as part of the sensitivity analysis.

#### Haematological safety

The presence of clinically significant haemolysis (≥25% haemoglobin reduction from baseline to <7 g/dL) between days 1–14 was described. Multivariable linear regression was used to model the maximum haemoglobin drop from baseline over 14 days, including as covariates: daily primaquine dose, baseline haemoglobin, log_2_ G6PD activity, age, sex, baseline parasite density, study site, and a dose-sex-log_2_ G6PD interaction. A DAG guided model specification (**Figure S*4***), and splines were used to assess nonlinear effects. Additional analyses, including a subgroup analysis of incident anaemia and analysis of methaemoglobinaemia, are described in Supplementary Material (**List S*2***).

Risk of bias was assessed using the ROB2 (randomised trials)^21^ and ROBINS-I tools (non-randomised studies),^22^ adapted for the current study objectives. Statistical analysis was performed using R (v4.3.0) according to a pre-specified plan.^23^ We conducted posthoc analyses presented in Discussion, including an aggregate meta-analysis of trials directly comparing high and low total doses outside Indonesia, to strengthen our findings.

## Results

Between 1 January 2000 and 23 July 2024, 234 *P. vivax* efficacy studies were published, of which ten included Indonesian study sites and were eligible for the pooled analysis. IPD from eight studies were available for analysis. After patient-level exclusion criteria were applied, 1797 patients enrolled into seven studies were included in the analysis (**Table *1***). The seven studies included one trial that individually randomised patients to different primaquine dose regimens,^15^ while the remaining six administered the same total primaquine dose within each study (a target total of either 3.5 or 7 mg/kg).^11–14,24,25^ There was a low to moderate risk of within study bias (**Table S*2***).

**Table 1.** Patient characteristics by study.

| Characteristic | Study |  |  |  |  |  |  | Overall |
| --- | --- | --- | --- | --- | --- | --- | --- | --- |
|  | Hasugian et al.<br>2007<sup>24</sup> | Pasaribu et al.<br>2013<sup>14</sup> | Sutanto et al.<br>2013<sup>13</sup> | Lidia et al.<br>2015<sup>25</sup> | Nelwan et al.<br>2015<sup>11</sup> | Taylor et al.<br>2019<sup>15</sup> | Poespoprodjo et<br>al. 2022<sup>12</sup> |  |
| **Number of patients** | 139 | 331 | 39 | 51 | 118 | 963 | 156 | 1797 |
| **Median age (Q1, Q3; years)** | 13 (4, 26) | 14 (9, 27) | 27 (25, 29) | 33 (25, 41) | 28 (25, 31) | 16 (10, 29) | 16 (7, 29) | 17 (10, 29) |
| **Age group** |  |  |  |  |  |  |  |  |
| Less than 5 years | 44 (32%) | 26 (8%) | 0 (0%) | 0 (0%) | 0 (0%) | 69 (7%) | 33 (21%) | 172 (9%) |
| 5 to 14 years | 29 (21%) | 148 (45%) | 0 (0%) | 0 (0%) | 0 (0%) | 386 (40%) | 42 (27%) | 605 (34%) |
| At least 15 years | 66 (47%) | 157 (47%) | 39 (100%) | 51 (100%) | 118 (100%) | 508 (53%) | 81 (52%) | 1020 (57%) |
| **Male sex** | 73 (53%) | 186 (56%) | 39 (100%) | 24 (47%) | 118 (100%) | 534 (55%) | 82 (53%) | 1056 (59%) |
| **Body weight (kg)** | 43 (14, 53) | 38 (22, 52) | 64 (59, 72) | 42 (36, 45) | 69 (63, 74) | 44 (24, 55) | 45 (19, 56) | 45 (24, 57) |
| **Baseline parasite density (per μL)** | 1575 (375, 5244) | 760 (320, 3100) | 2848 (784, 4432) | 2640 (1500, 6560) | 880 (148, 2496) | 3911 (1170, 8659) | 4920 (1680, 11550) | 2793 (630, 7130) |
| **Presence or recent history of fever** | 139 (100%) | 321 (97%) | 6 (16%) | 51 (100%) | NA | 850 (88%) | 156 (100%) | 1523 (91%) |
| Number of missing data | 0 | 0 | 2 | 0 | 118 | 0 | 0 | 120 |
| **Schizonticidal drug** |  |  |  |  |  |  |  |  |
| Dihydroartemisinin-Piperaquine | 68 (49%) | 164 (50%) | 0 (0%) | 25 (49%) | 58 (49%) | 961 (100%) | 156 (100%) | 1432 (80%) |
| Artesunate-Amodiaquine | 71 (51%) | 167 (50%) | 0 (0%) | 0 (0%) | 0 (0%) | 0 (0%) | 0 (0%) | 238 (13%) |
| Artesunate-Pyronaridine | 0 (0%) | 0 (0%) | 0 (0%) | 0 (0%) | 60 (51%) | 0 (0%) | 0 (0%) | 60 (3.3%) |
| Chloroquine | 0 (0%) | 0 (0%) | 0 (0%) | 26 (51%) | 0 (0%) | 2 (0.2%) | 0 (0%) | 28 (1.6%) |
| Quinine | 0 (0%) | 0 (0%) | 39 (100%) | 0 (0%) | 0 (0%) | 0 (0%) | 0 (0%) | 39 (2.2%) |
| **Primaquine total dose (mg base/kg)** | 3.8 (3.4, 4.3) | 4.2 (3.6, 4.8) | 7.1 (6.7, 8.2) | 4.5 (4.2, 4.9) | 7.1 (6.5, 8.1) | 7.0 (6.0, 7.9) | 7.3 (5.9, 8.2) | 6.4 (4.1, 7.5) |
| **Primaquine total dose group** |  |  |  |  |  |  |  |  |
| No primaquine (0 mg base/kg) | 0 (0%) | 0 (0%) | 0 (0%) | 0 (0%) | 0 (0%) | 196 (20%) | 0 (0%) | 196 (11%) |
| Low ($\geq 2$ and $< 5$ mg base/kg) | 118 (85%) | 265 (80%) | 0 (0%) | 39 (76%) | 1 (0.85%) | 21 (2.2%) | 20 (13%) | 464 (26%) |
| High ($\geq 5$ mg base/kg) | 21 (15%) | 66 (20%) | 39 (100%) | 12 (24%) | 117 (99%) | 746 (77%) | 136 (87%) | 1137 (63%) |
| **Primaquine daily dose (mg base/kg)** | 0.27 (0.24, 0.31) | 0.30 (0.26, 0.34) | 0.51 (0.48, 0.58) | 0.32 (0.30, 0.35) | 0.53 (0.47, 0.61) | 0.59 (0.46, 1.00) | 0.43 (0.00, 0.56) | 0.47 (0.28, 0.63) |
| **Primaquine duration** |  |  |  |  |  |  |  |  |
| No primaquine | 0 (0%) | 0 (0%) | 0 (0%) | 0 (0%) | 0 (0%) | 196 (20%) | 0 (0%) | 196 (11%) |
| 7 days | 0 (0%) | 0 (0%) | 0 (0%) | 0 (0%) | 0 (0%) | 385 (40%) | 0 (0%) | 385 (21%) |
| 14 days | 139 (100%) | 331 (100%) | 39 (100%) | 51 (100%) | 118 (100%) | 382 (40%) | 156 (100%) | 1216 (68%) |
| **Primaquine dose calculation** |  |  |  |  |  |  |  |  |
| No primaquine | 0 (0%) | 0 (0%) | 0 (0%) | 0 (0%) | 0 (0%) | 196 (20%) | 0 (0%) | 196 (11%) |
| Actual dosing | 0 (0%) | 0 (0%) | 38 (97%) | 0 (0%) | 118 (100%) | 767 (80%) | 156 (100%) | 1079 (60%) |
| Protocol dosing | 139 (100%) | 331 (100%) | 1 (2.6%) | 51 (100%) | 0 (0%) | 0 (0%) | 0 (0%) | 522 (29%) |
| **Primaquine supervision** |  |  |  |  |  |  |  |  |
| No primaquine | 0 (0%) | 0 (0%) | 0 (0%) | 0 (0%) | 0 (0%) | 196 (20%) | 0 (0%) | 196 (11%) |
| Unsupervised | 139 (100%) | 0 (0%) | 0 (0%) | 0 (0%) | 0 (0%) | 0 (0%) | 67 (43%) | 206 (11%) |
| Partially supervised | 0 (0%) | 0 (0%) | 0 (0%) | 0 (0%) | 0 (0%) | 0 (0%) | 89 (57%) | 89 (5.0%) |
| Fully supervised | 0 (0%) | 331 (100%) | 39 (100%) | 51 (100%) | 118 (100%) | 767 (80%) | 0 (0%) | 1306 (73%) |
| **Baseline haemoglobin level (g/dL)** | 11·0 (8·8, 12·4) | 11·9 (10·8, 12·8) | 14·3 (13·1, 14·7) | 10·3 (9·5, 11·2) | 13·8 (13·0, 14·5) | 12·6 (11·4, 13·9) | 11·6 (10·4, 13·1) | 12·3 (11·0, 13·7) |
| Number of missing data | 2 | 0 | 0 | 0 | 0 | 0 | 0 | 2 |
| **Day 7 methaemoglobin level (%)** | NA | 3·8 (2·7, 5·7) | 5·6 (4·1, 7·1) | NA | 5·8 (3·7, 8·2) | 8·3 (5·4, 12) | NA | 6·0 (3·5, 9·4) |
| Number of missing data | 139 | 28 | 1 | 51 | 0 | 536 | 156 | 911 |
| **Transmission intensity** |  |  |  |  |  |  |  |  |
| Low | 0 (0%) | 0 (0%) | 0 (0%) | 0 (0%) | 0 (0%) | 0 (0%) | 0 (0%) | 0 (0%) |
| Moderate | 0 (0%) | 331 (100%) | 0 (0%) | 0 (0%) | 0 (0%) | 963 (100%) | 156 (100%) | 1450 (81%) |
| High | 139 (100%) | 0 (0%) | 39 (100%) | 51 (100%) | 118 (100%) | 0 (0%) | 0 (0%) | 347 (19%) |
| **Origin of infections** |  |  |  |  |  |  |  |  |
| Within Papua | 139 (100%) | 0 (0%) | 39 (100%) | 0 (0%) | 118 (100%) | 0 (0%) | 156 (100%) | 452 (25%) |
| Outside Papua | 0 (0%) | 331 (100%) | 0 (0%) | 51 (100%) | 0 (0%) | 963 (100%) | 0 (0%) | 1345 (75%) |
| **Study duration** |  |  |  |  |  |  |  |  |
| At least 180 days | 0 (0%) | 331 (100%) | 39 (100%) | 0 (0%) | 118 (100%) | 963 (100%) | 156 (100%) | 1607 (89%) |
| Less than 180 days | 139 (100%) | 0 (0%) | 0 (0%) | 51 (100%) | 0 (0%) | 0 (0%) | 0 (0%) | 190 (11%) |

Numbers are in median (first quartile [Q1], third quartile [Q3]) or frequency (percentage). NA, not available; kg, kilogram; µL, microlitre; g, gram; dL, decilitre; mg, milligram. Transmission intensity was categorised as low (<1 case per 1000 person-years), moderate (1 case to <10 cases per 1000 person-years), and high (≥10 cases per 1000 person-years) according to subnational malaria incidence estimates for the median year of study enrolment.<sup>49</sup> Patients contributing to the tolerability and safety datasets, as well as any sensitivity or restricted analyses, were subsets of the dataset shown in this table.

The median age of the patients was 17 years (interquartile range [IQR] 10–29) with 172 (9·6%) younger than 5 years. The median weight was 45 kg (IQR 24–57) and 1056 (59%) were male. Overall, 452 (25%) patients acquired their *P. vivax* infection in Papua (**Figure *2***) and 1432 (80%) were treated with dihydroartemisinin-piperaquine (**Table *1***). A total of 196 (11%) patients were treated without primaquine, 464 (26%) with low total dose, and 1137 (63%) with high total dose (**Figures S*5*, S*6*, S*7*, S*8*).**

**Figure 1.**
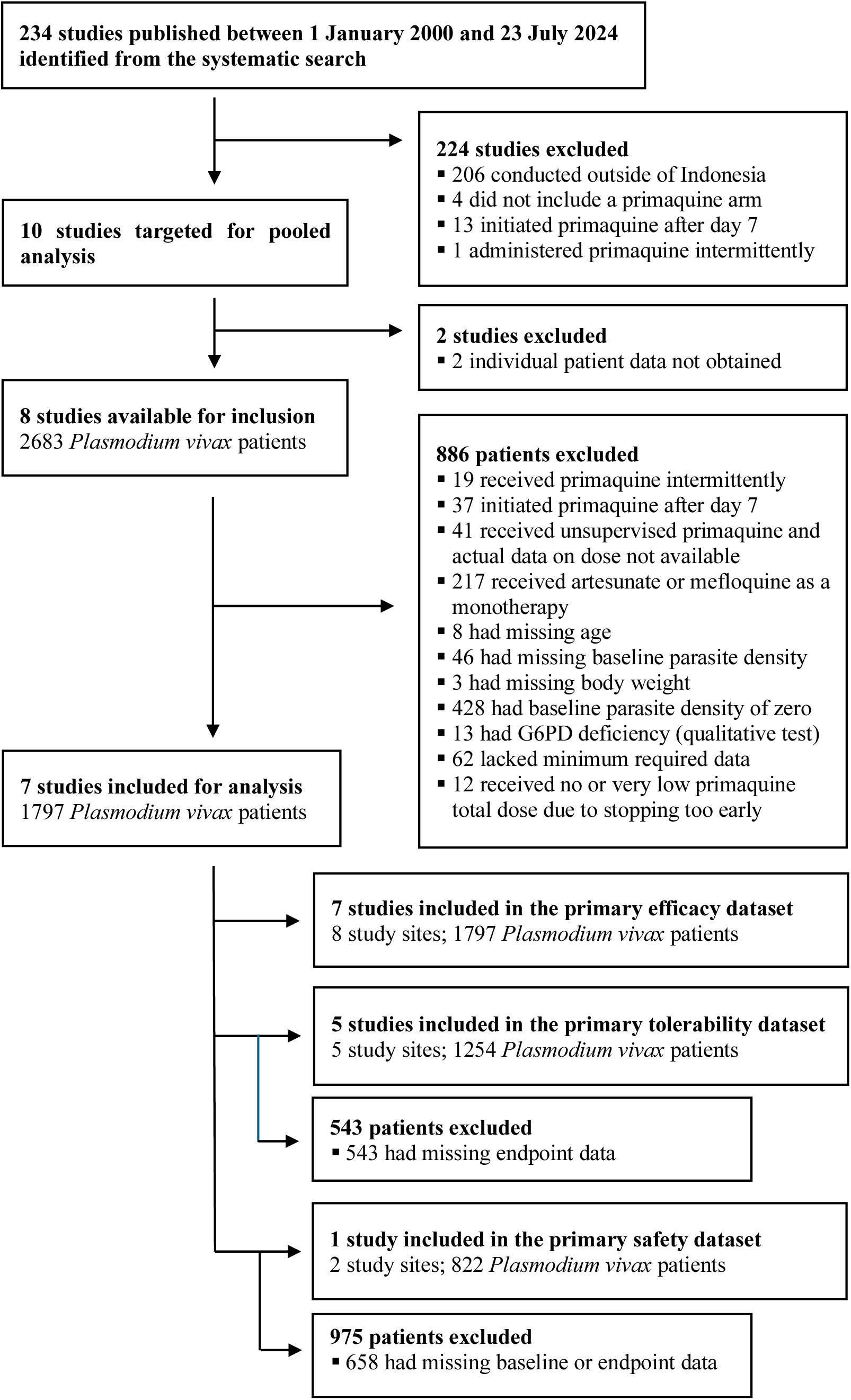
Study and patient selection. Databases systematically searched included MEDLINE, Embase, Web of Science, Scopus and the Cochrane Library. Additional analyses (e.g., sensitivity or restricted analyses) were conducted on smaller subsets of the relevant primary dataset. Forty-one patients were excluded as protocol-based dose calculation was considered unreliable in the context of unsupervised primaquine. Missing endpoint data in the tolerability and safety datasets primarily reflects studies that did not systematically measure the outcome of interest.

**Figure 2.**
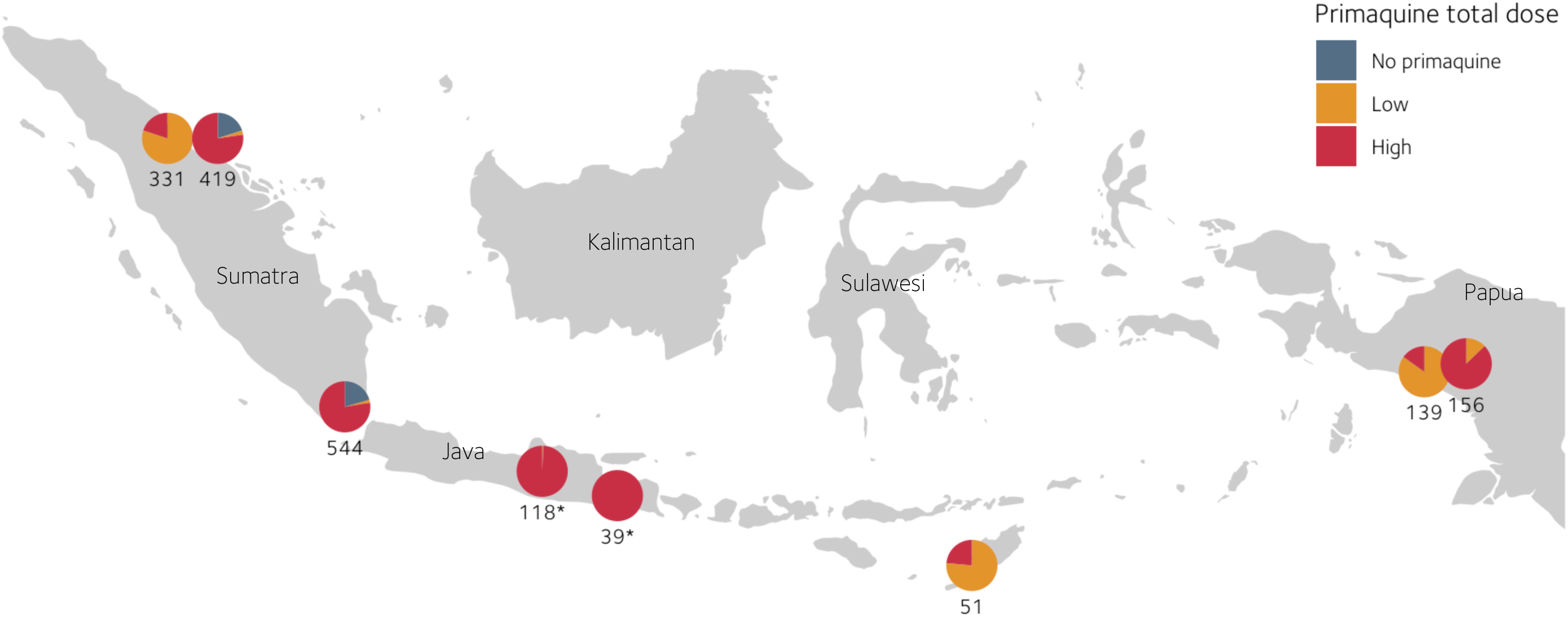
Study sites that contributed to the pooled individual patient data across Indonesia. Each circle represents a study site, and the number of participants included in the pooled data. Primaquine total dose: no primaquine (0 mg/kg), low (≥2 and <5 mg/kg), high (≥5 mg/kg). Basemap shapefile data were obtained from the publicly available Natural Earth project (https://www.naturalearthdata.com) and accessed via the open-source R packages maps^50^ and ggplot2.^51^ * Malaria-naive soldier patients were deployed and acquired infections in Papua then returned to malaria-free Java for follow-up (also referred to as the soldier relapse model^11,13^). For a more detailed description of the study sites, please refer to Table S4.

Compared with low total dose, patients administered high-dose primaquine were more likely to be treated over seven days, receive dihydroartemisinin-piperaquine, have a higher baseline parasite density, and have actual dosing data available (**Table S*3***, **Figure S*9***). Eligible studies that were not included^26–28^ predominantly enrolled patients in Papua and had a shorter follow-up duration (**Tables S*4*, S*5*, S*6***).

Compared with patients treated without primaquine (enrolled in a single study conducted at non-Papuan sites), the rate of *P. vivax* recurrence by day 180 was lower in both patients treated with low-dose (3·5 mg/kg) primaquine (adjusted hazard ratio [AHR] 0·33; 95% CI 0·25 to 0·44) and high-dose (7 mg/kg) primaquine (AHR 0·18; 95% CI 0·12 to 0·26). The corresponding rate of recurrence was lower following high-dose primaquine compared to low-dose primaquine (AHR 0·53; 95% CI 0·45 to 0·63; **Figure *3***). Increasing the total dose above 7 mg/kg remains of uncertain efficacy as the interval estimates of the estimated dose-response curve were wide (**Figure *3***). This dose-response relationship was consistent across alternative dose groupings and sensitivity analyses (**Figures S*11***, **S*12***). Negative control methods suggest residual confounding was minimal (**List S*3***). The effect of increasing the dose of primaquine did not differ with age (<5 or ≥5 years old, p_interaction_ = 0·88) or by origin of infection (within Papua or outside Papua, p_interaction_ = 0·45, **Figure S*13***).

**Figure 3.**
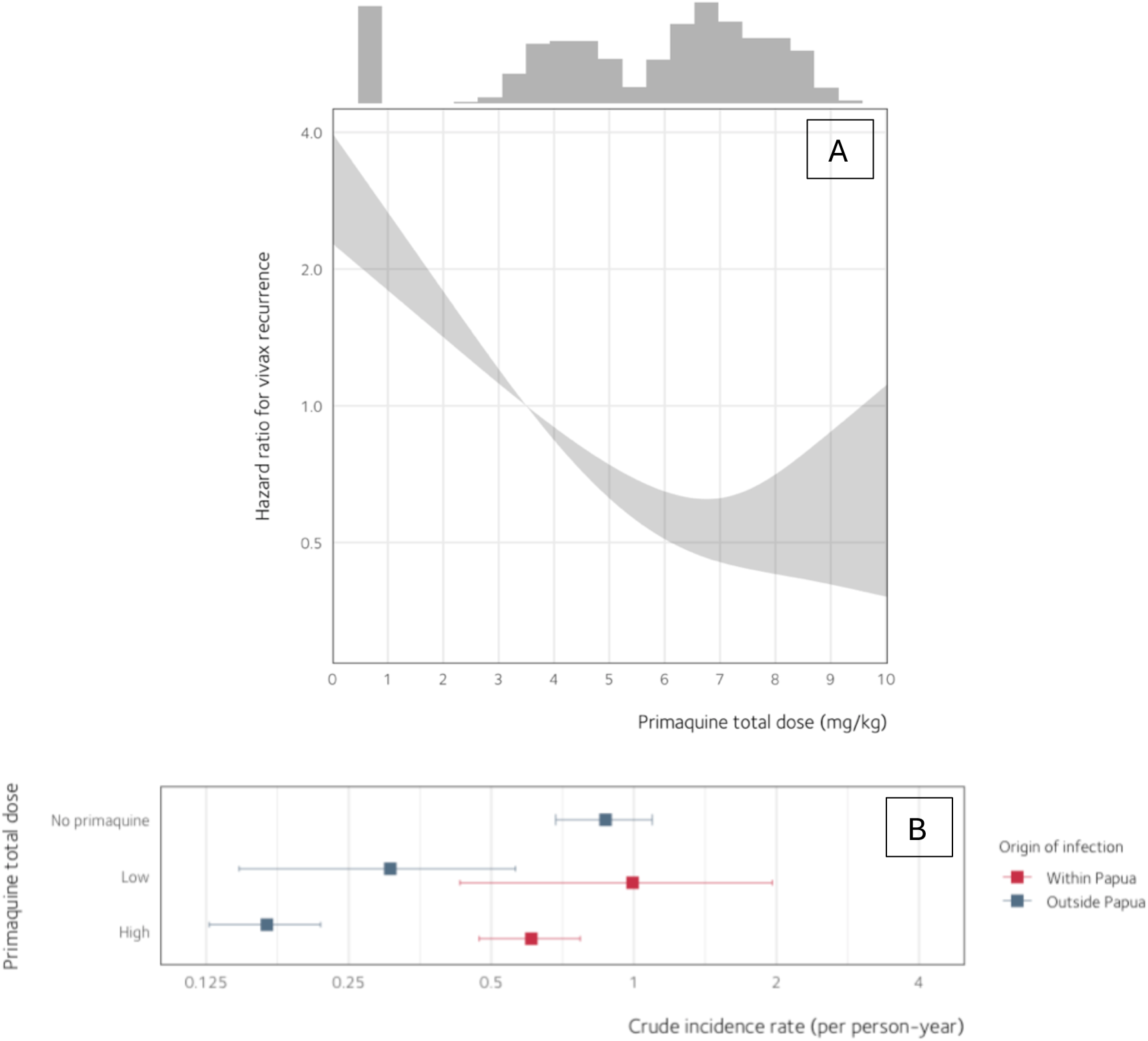
Effect of increasing primaquine total dose on the rate of *P. vivax* recurrence by day 180. (A) The reference value (HR = 1) was set at the target low dose of 3·5 mg/kg. The shaded region shows 95% CI. Estimates were derived from a multivariable Cox proportional hazards model, fitted to the primary efficacy dataset (n = 1797). The histogram along the top margin shows the distribution of primaquine total doses, with the leftmost bar representing patients who were treated without primaquine (i.e., 0 mg/kg). The vertical axis is shown on a logarithmic scale. (B) Crude incidence rates in the subset of patients (n = 1607) followed for up to 180 days across multiple *P. vivax* episodes by primaquine total dose and origin of infection. Patients treated without primaquine were only available from outside Papua. Point estimates (solid squares) were calculated as the number of *P. vivax* episodes divided by person-years at risk, with 95% CI for a Poisson rate obtained using an exact method. The horizontal axis is shown on a logarithmic scale. For a more detailed description of the study sites and the regression output, please refer to Tables S4 and S7, respectively.

In a subset of patients (n = 1608, 89%) followed through multiple P. vivax episodes for at least 180 days, those who acquired infections within Papua had a higher 180-day recurrence rate (76 episodes per 120 person-years; crude incidence rate [IR] 0·63 per person-year; 95% confidence interval [CI] 0·50 to 0·79) than those infected outside Papua (140 episodes per 451 person-years; IR 0·31 per person-year; 95% CI 0·26 to 0·37). **Figure *3*** shows recurrence rates for both regions by primaquine dose groupings.

Gastrointestinal tolerability on days 5–7 was assessed in 1254 (70%) patients from five studies.^12,14,15,24,25^ Of these, 190 (15%) received no primaquine, 203 (16%) received low daily dose, 482 (38%) intermediate daily dose, and 379 (30%) high daily dose primaquine; with 125 (10%) younger than 5 years. Within each primaquine daily dose group, there was substantial heterogeneity in the percentage of patients reporting gastrointestinal discomfort on days 5–7 across studies (**Figure S*14*, Table S*7***), making data pooling for effect estimation unreliable. To estimate the unbiased effect of increasing daily doses, we used subsets of data from one trial^15^ in which primaquine daily doses were randomised. The risk of early gastrointestinal symptoms (day 0 and days 1–2) did not vary with the daily dose of primaquine (**Figure S*15*, Table S*9***). However, by days 5–7, when 894/952 (94%) patients were afebrile, the risk of gastrointestinal symptoms increased with the daily dose of primaquine: adjusted risk ratio [ARR] 1·32 (95% CI 1·15 to 1·51) per 0·25 mg/kg/day increment in primaquine dose (**Figure S*15*, Table S*9***). In 767 patients assessable for acute vomiting, there was inconclusive evidence that a higher daily dose of primaquine resulted in an increased risk of acute vomiting within 60 minutes of administration (ARR 1·19; 95% CI 0·84 to 1·67 per 0·25 mg/kg/day increment; **Figure S*16*, Table S8**).

One multi-country randomised trial^15^ contributed 822 patients to the primary haematological safety analysis, of whom 173 (21%) received no primaquine, 6 (<1%) received a low daily dose, 318 (39%) received an intermediate daily dose, and 325 (40%) a high daily dose. G6PD activity was measured at baseline in all eligible patients and was similar between treatment arms; 788 (96%) had normal G6PD activity (≥70%) and 34 (4%) had intermediate activity (30–69%; **Figure S*17***).

Severe haemolysis was recorded in one patient, a female in the 5–14-year age group with 37·4% G6PD activity treated with dihydroartemisinin-piperaquine plus a high daily dose of primaquine (0·99 mg base/kg) administered over 7 days. She had a 41% fall in haemoglobin from 11·6 g/dL on day 0 to 6·9 g/dL on day 3 (4·7 g/dL reduction) and recovered (Hb ≥11 g/dL) at least by day 21 without requiring a blood transfusion. No patients had a haemoglobin fall to <5 g/dL, a haemoglobin fall >5 g/dL from baseline, blood transfusion, renal failure requiring dialysis, or death (**Table S*10***).

The effect of primaquine daily dose on haemoglobin varied with sex and G6PD activity (p_interaction_ = 0·01). In contrast to females with normal G6PD activity, a higher daily dose of primaquine was associated with a greater absolute fall in haemoglobin in females with intermediate G6PD activity (**Figures *4*A, S*18***). In males, lower G6PD activity was not associated with a greater absolute decrease in haemoglobin. In 612 patients with normal G6PD activity and a baseline haemoglobin level of ≥11 g/dL, the risk of anaemia on days 2–3 did not increase with higher primaquine daily doses (**Figure S*19***, **Table S*9***). In 788 patients with normal G6PD activity, haemoglobin levels reached an expected nadir on days 2–3, likely reflecting haemolysis associated with blood-stage acute malaria. Haemoglobin levels returned to baseline approximately two weeks after initiating treatment (**Figure *4*B**). A higher primaquine daily dose resulted in higher day 7 methaemoglobin levels and an increased risk of methaemoglobin levels ≥10% between days 1 to 14 (prevalence = 262/1026 [25·5%], **Figure S*20***).

**Figure 4.**
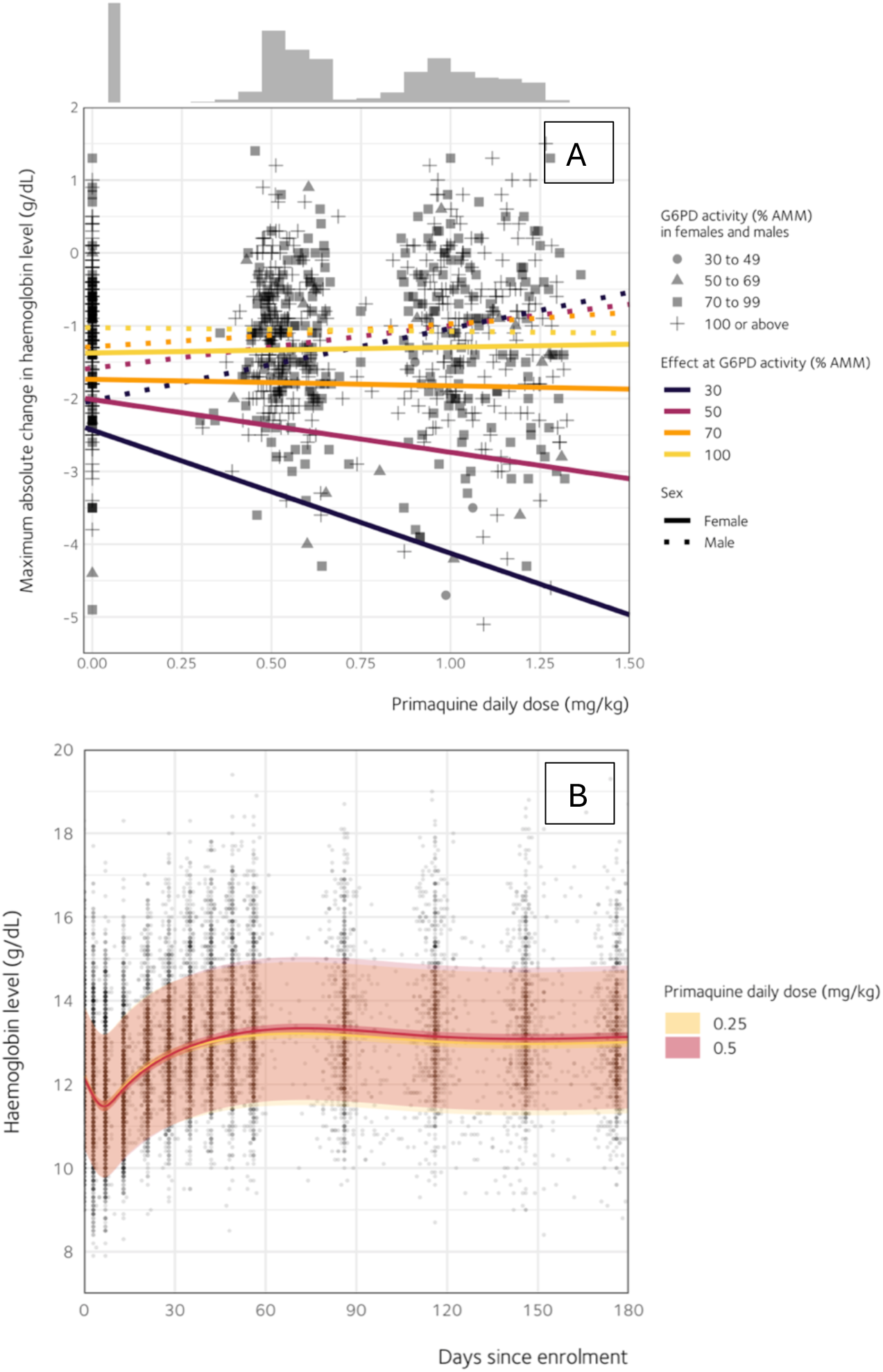
Effect of primaquine daily dose on haematological profile. (A) Maximum absolute change from baseline in haemoglobin levels between days 1 and 14 across primaquine daily doses, sex, and G6PD activity levels. Estimates were derived from a multivariable linear model fitted to the primary safety dataset (n = 822, 34 patients with intermediate G6PD activity between 30% and 69%). The plot shows model-implied predictions at four selected cut-offs (30%, 50%, 70%, and 100%) of G6PD activity to aid interpretation. The histogram along the top margin shows the distribution of primaquine daily doses in the model data, with the leftmost bar representing patients who were treated without primaquine (i.e., 0 mg/kg). (B) Temporal dynamics of haematological recovery in patients with G6PD activity of at least 70%. Estimates were derived from a multilevel, multivariable linear model (n = 790). Model-implied predictions at primaquine daily doses of 0·25 mg/kg and 0·5 mg/kg are shown. Solid curves represent the expected haemoglobin levels over time following treatment, with thick and thin shaded regions denoting the 95% confidence and prediction intervals, respectively. AMM, adjusted male median. For a more detailed description of the regression output, please refer to Table S7.

## Discussion

The optimal anti-relapse regimen to prevent recurrent *P. vivax* malaria must trade-off the absolute number of recurrent malaria episodes averted against the length of treatment duration, its tolerability, and safety. This balance of practicality, tolerability, and safety against absolute benefit will vary across regions primarily because the absolute risk of recurrence varies considerably. Our meta-analysis of 1797 Indonesian patients demonstrates that increasing the total dose of primaquine from 3·5 to 7 mg/kg would halve the recurrence rate. However, the absolute benefit provided by the higher dose will vary substantially as the crude recurrence incidence rate varied substantially between regions (IR 0·63 [95% CI 0·50 to 0·79] per person-year within Papua; IR 0·31 [95% CI 0·26 to 0·37] per person-year outside Papua). Higher daily doses were associated with a greater risk of gastrointestinal discomfort and there is a risk of severe primaquine-induced haemolysis in females with intermediate G6PD activity.^29^ Identifying an optimal balance between benefits and harms is key to determining the best regimen for each region within Indonesia.

Our Indonesia-specific estimates for anti-relapse efficacy comparing high- and low-dose primaquine align with those reported by Commons and colleagues using pooled global data.^6^ Both analyses included non-randomised comparisons, so confounding bias cannot be excluded. One limitation of our study is that all patients included in the no-primaquine group were from sites outside Papua, and there was no direct randomisation to the low-dose group. Much of the within-study dose variation was attributable to differences in body weight rather than to random allocation. Similar to Commons et al.,^6^ we used a causal diagram to inform adjustment for key confounders in regression analysis. We further applied negative control methods^30^ and found no material residual confounding. Consistent with this study, a meta-analysis of study-level data from randomised trials^31–35^ directly comparing high- and low-dose primaquine in any country supported that high-dose primaquine reduces the rate of first *P. vivax* recurrence by around 50% (**Figures *5*A, 5B**).

**Figure 5.**
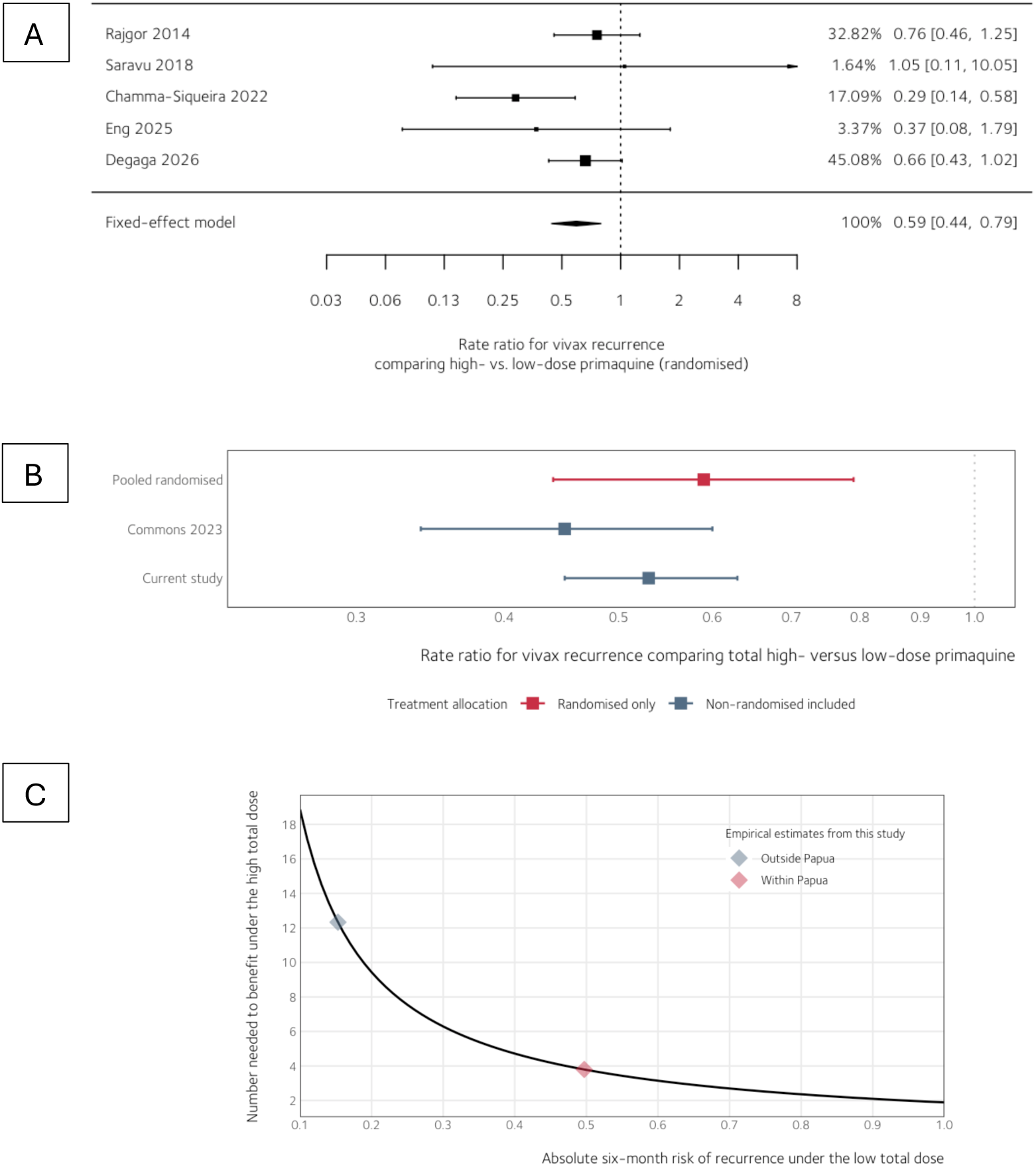
Meta-analysis of randomised controlled trials and its comparison to global and Indonesian IPD meta-analyses on the effect of high-dose versus low-dose primaquine. (A) Systematic search for trials that randomised high- and low-dose primaquine was conducted without country restrictions from 1960 to 2025 with a minimum follow-up of approximately six months. Estimates from the first two studies^31,32^ were derived from the number of events and the period at risk reported in each study to calculate the incidence rate ratio (patients censored before the end of the study were assumed to contribute half of their respective follow-up period). Estimates from the last three studies^33–35^ were obtained directly from the hazard ratio reported in each study. In the current context, these effect measures (i.e., the incidence rate ratio and hazard ratio) were assumed to be approximately interchangeable and summarised here as the rate ratio. Vertical dashed line indicates an equal effect (RR = 1). Estimates to the left of the dashed line indicate greater efficacy of high dose primaquine. Estimates to the right indicate greater efficacy of low dose primaquine. (B) Pooled randomised estimates were derived from a study-level global meta-analysis presented in the top panel. Estimates from Commons 2023^6^ were derived from an IPD meta-analysis of randomised and non-randomised *P. vivax* efficacy studies conducted globally. Estimates from the current study were derived by comparing the hazard ratio estimate at 7 mg/kg with that at 3·5 mg/kg in the Indonesian pooled data. The solid square represents a point estimate with a 95% confidence interval. The horizontal axis is shown on a logarithmic scale. CI, confidence interval. IPD, individual patient data. (C) Expected number needed to benefit with total high-dose primaquine (compared with total low-dose primaquine, assuming a risk ratio of 0.53) across varying absolute risks of recurrence over a 6-month period.

With intense, year-round malaria transmission, infections in Papua may involve a higher hypnozoite burden, as well as, ’Chesson-like’ strains that are more tolerant to primaquine, both necessitating a higher primaquine dose.^36–38^ In contrast, regions outside Papua, such as Sumatra, experience low to moderate and increasingly focal transmission approaching elimination, where residual burden is often linked to mobile and migrant populations and *P. vivax* predominance.^39^ Lower force of reinfection means observed recurrences may be more likely to reflect hypnozoite activation, and treatment responses could therefore appear more favourable than in Papua. However, we found no evidence that higher doses of primaquine in patients infected in Papua would provide greater relative efficacy compared to patients infected outside Papua. The global analysis reported similar findings, showing high relative efficacy of high-dose primaquine in locations with both low- and high-relapse periodicity.^6^ Hence, higher relative anti-relapse efficacy of increased primaquine doses is likely consistent and generalisable across different regions and parasite strains. However, given the diverse case-mix and varying baseline risks across populations, heterogeneous treatment-effects on an absolute scale are expected and will be relevant for policy makers and clinicians. When comparing high versus low total-dose primaquine, the number needed to benefit to prevent one recurrence over six months in Papua would be around four patients, compared with about twelve patients in non-Papua regions (**Figure *5*C**).

A higher daily dose of primaquine led to increased gastrointestinal discomfort, independent of acute malaria symptoms. Reduced tolerability could lead to non-compliance, preventing the target total dose from being achieved. Reduced adherence or inadequate supervision may lead to lower anti-relapse efficacy.^12,40^ Although, previous studies have shown that taking primaquine tablets with a meal significantly improves gastrointestinal tolerability,^41^ relevant data to address this in our IPD analysis were not available.^42^

A lack of routine G6PD screening limits the safe implementation of high total dose primaquine. The risk of haemolysis is almost entirely in individuals with G6PD deficiency. While our analysis did not include this population, our analysis included patients with intermediate G6PD activity. Chu and colleagues reported that despite being labelled as phenotypically normal by point-of-care tests (≥30% G6PD activity), G6PD heterozygous females can experience significant haemolysis when exposed to 1 mg/kg/day of primaquine.^29^ We observed that females with G6PD activity between 30 and 69% experienced greater haemoglobin reductions as the daily dose increased compared to those with G6PD activity ≥70%, however this was not clinically significant at doses of 0.5 mg/kg/day or less. Primaquine also induced dose-dependent increases in methaemoglobin levels in our study population. Prior evidence suggests that this elevation typically occurs without symptoms, and resolves spontaneously within weeks.^41,43^ Increased primaquine-induced methaemoglobinaemia indicates sufficient production of active primaquine metabolites and hence, greater anti-relapse efficacy.^44,45^

In line with the 2024 WHO antimalarial guidelines,^8^ our findings suggest that for most patients with *P. vivax* in Indonesia, an optimal regimen could involve a high total dose of 7 mg/kg administered over 1–2 weeks, taken with food, and initiated after screening for G6PD deficiency. Of note, in western Indonesia, there is a low risk of relapse following a low total dose primaquine,^14^ suggesting that the absolute benefit from high total dose primaquine may be reduced in these areas and different dosing could be considered. A pragmatic next step could be a tiered national strategy aligned to administrative boundaries, with 7 mg/kg prioritised in higher-relapse settings and lower doses considered in lower-risk areas, supported by robust surveillance of recurrence and safety outcomes. Mathematical scenario modelling could further inform optimal doses under varying local transmission and programmatic conditions.

Tafenoquine has been recommended for use in endemic countries in South America, where chloroquine is typically the blood-stage drug of choice.^8^ This single-dose anti-relapse treatment is currently not recommended for patients receiving artemisinin-based combination therapies (ACTs).^8^ In Indonesia, where ACTs are recommended due to a high prevalence of chloroquine-resistant *P. vivax*, tafenoquine is not currently available. The INSPECTOR trial, conducted in soldiers returning from Indonesian Papua, found no clinically meaningful benefit of tafenoquine plus dihydroartemisinin-piperaquine compared with dihydroartemisinin-piperaquine alone.^26^ The reason for this remains unclear, although a drug–drug interaction or suboptimal dosing have been proposed.^46^ A randomised pharmacokinetic trial found no evidence of reductions in tafenoquine exposure when co-administered with dihydroartemisinin-piperaquine.^47^ Ongoing trials (e.g., TADORE+ [NCT07060794], SEADOT [NCT04704999]) will further assess potential drug–drug interactions and whether suboptimal dosing may account for the observed discrepancies.

We were unable to include two recently published eligible studies in our pooled dataset, primarily due to unavailability of IPD. These studies would have added <10% more patients, making it unlikely that the current results would change significantly. One of these studies^26^ randomised patients to receive low-dose primaquine over 14 days versus a placebo, and its estimated efficacy (HR 0·26; 95% CI 0·16 to 0·43) aligns with our pooled effect-estimates. In the current study, ∼30% of patients exposed to primaquine were estimated from the study protocol rather than the actual tablets administered. Given an inability to account for reduced adherence this may have led to the actual dose of primaquine received being less than expected, which would be expected to bias the anti-relapse efficacy towards the null, making our efficacy estimates conservative. While the same applies to tolerability and safety (i.e., higher daily doses potentially appearing more tolerable and safer), both in practice can be effectively mitigated by taking food and G6PD screening. Therefore, the impact of potential quantitative bias is deemed minimal for clinical use. Although our focus was on anti-relapse efficacy, the studies (except for those conducting follow-up in malaria-free regions^11,13^) were unable to distinguish whether recurrences were caused by new infections, treatment failure, or hypnozoite activation. The use of highly efficacious schizonticides made recrudescence unlikely and approximately 80% of vivax recurrences were estimated to result from relapses.^48^ If new infections were indeed substantial, they would have diluted our efficacy estimates towards the null, making our results conservative. Lastly, while underlying transmission intensity could confound the estimated causal effect and may vary across locations and over time, transmission levels were broadly stable within sites across the years. We therefore adjusted for study site to account for spatial and temporal differences, and a negative outcome control analysis using *P. falciparum* infection suggested that any residual confounding was minimal.

In summary, increasing the total primaquine dose from 3·5 mg/kg to 7 mg/kg is likely to provide a substantial reduction in the risk of recurrence, with no evidence of treatment-effect heterogeneity across Indonesia. However, the variable risk of recurrence across the country means that there are substantial differences in the absolute benefit of 7 mg/kg versus 3·5 mg/kg total dose primaquine (e.g., higher benefit in Papua but lower in Sumatra). Completing a high total dose of primaquine in a shorter timeframe by administering a higher daily dose, is preferable for maximising adherence to a full course of treatment, however this may increase the risk of gastrointestinal discomfort or haemolysis. These risks can be mitigated by co-administering primaquine with food, G6PD screening, and monitoring for potential haemolytic events. Future efforts should prioritise optimising implementation, adherence, and safe delivery of the regimens rather than pursuing evaluation of higher primaquine total doses above 7 mg/kg.

## Contributors

IF, JAW, RNP, JKB, and RJC conceived the study, analysed and interpreted the data, and drafted the manuscript. IF and RJC accessed and verified the data. JAW, RNP, and JKB provided technical support. APP, IS, EJN, KL, IRFE, WRJT, KT, NPJD, JRP, JAS, RNP, JKB conceived and undertook the individual studies and enrolled the patients. All authors revised the manuscript and were responsible for the decision to submit for publication.

## Data sharing

Pseudo-anonymised participant data used in this analysis are available for access via the Infectious Diseases Data Observatory (IDDO) website (https://www.iddo.org). Requests for data access will be reviewed according to IDDO procedures, to ensure that use of data protects the interests of the participants and researchers according to the terms of ethics approval and principles of equitable data sharing. IDDO is registered with the Registry of Research Data Repositories (https://www.re3data.org/). Code for data analysis and visualisation is available at https://github.com/ihsanfadil/wwarn-pq-ina.

## Data Availability

Pseudo-anonymised participant data used in this analysis are available for access via the Infectious Diseases Data Observatory (IDDO) website (https://www.iddo.org). Requests for data access will be reviewed according to IDDO procedures, to ensure that use of data protects the interests of the participants and researchers according to the terms of ethics approval and principles of equitable data sharing. IDDO is registered with the Registry of Research Data Repositories (https://www.re3data.org/).

## Acknowledgements

IF is supported by the Nuffield Department of Medicine Tropical Network Fund. RJC, RNP, JAS and KT are supported by the Australian National Health and Medical Research Council (NHMRC) Investigator Grants (1194702, 2008501, 1196068 and 2033264 respectively). JAW is a Sir Henry Dale Fellow funded by the Wellcome Trust (223253/Z/21/Z). MR is supported by the Australian NHMRC CRE Grant 2024622 (Australian Centre of Research Excellence for Malaria Elimination) and Australian NHMRC Synergy Grant 2018654. We thank all patient volunteers, healthcare workers, and research staff who contributed to the individual studies at all the sites, and the WWARN team for technical and administrative support. This research was supported by grants from Bill and Melinda Gates Foundation. For open access, the authors have applied a CC BY public copyright licence to any Author Accepted Manuscript arising from this submission. The findings and conclusions in this report are those of the authors and do not necessarily represent the official position of the US Centers for Disease Control and Prevention.

## Declaration of generative AI and AI-assisted technologies in the manuscript preparation process

During the preparation of this work the author(s) used GPT-5 to perform spell-checking and review computer code for errors. After using this tool/service, the author(s) reviewed and edited the content as needed and take(s) full responsibility for the content of the published article.

## Supplementary Material

### List S1. Systematic search terms for the databases

Vivax AND (artefenomel OR arterolane OR amodiaquine OR atovaquone OR artemisinin OR arteether OR artesunate OR artemether OR artemotil OR azithromycin OR artekin OR chloroquine OR chlorproguanil OR cycloguanil OR clindamycin OR coartem OR dapsone OR dihydroartemisinin OR duo-cotecxin OR doxycycline OR halofantrine OR lumefantrine OR lariam OR malarone OR mefloquine OR naphthoquine OR naphthoquinone OR piperaquine OR primaquine OR proguanil OR pyrimethamine OR pyronaridine OR proguanil OR quinidine OR quinine OR riamet OR sulphadoxine OR tetracycline OR tafenoquine)

### List S2. Supplementary methods

#### Additional inclusion criteria

For the haematological safety analysis, only patients with G6PD activity ≥30% of the adjusted male median (AMM) or a negative qualitative test were included. Reporting of parasite presence or absence during follow-up, presence of gastrointestinal discomfort (vomiting, anorexia, diarrhoea) during follow-up, and haemoglobin measured on day 0 and at least once during follow up were required for anti-relapse efficacy, gastrointestinal tolerability, and haematological safety analyses, respectively. For Poespoprodjo 2022,^12^ we included only patients for whom actual dose data were available. Protocol-based dose calculation was considered unreliable in the context of unsupervised primaquine.

#### Categories of total and daily primaquine doses

| Category | Body weight-adjusted primaquine exposure (mg/kg) |  |
| --- | --- | --- |
|  | Daily | Total |
| No primaquine | 0 | 0 |
| Very low | ... | $>0$ and $<2$ |
| Low | $>0$ and $<0.375$ | $\geq 2$ and $<5$ |
| Intermediate | $\geq 0.375$ and $<0.75$ | ... |
| High | $\geq 0.75$ | $\geq 5$ |

#### Follow-up periods of the primary endpoints

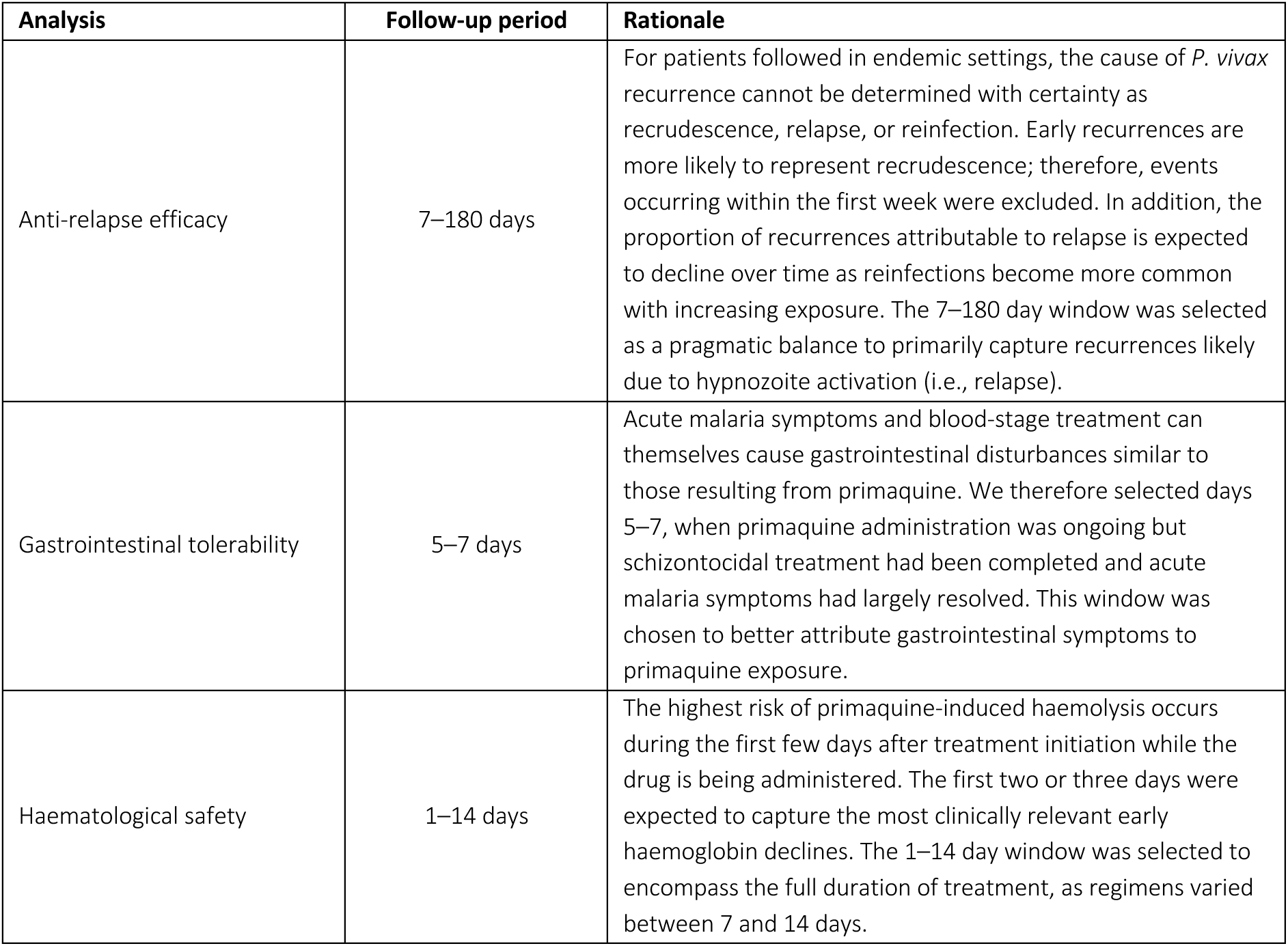

| Analysis | Follow-up period | Rationale |
| --- | --- | --- |
| Anti-relapse efficacy | 7–180 days | For patients followed in endemic settings, the cause of *P. vivax* recurrence cannot be determined with certainty as recrudescence, relapse, or reinfection. Early recurrences are more likely to represent recrudescence; therefore, events occurring within the first week were excluded. In addition, the proportion of recurrences attributable to relapse is expected to decline over time as reinfections become more common with increasing exposure. The 7–180 day window was selected as a pragmatic balance to primarily capture recurrences likely due to hypnozoite activation (i.e., relapse). |
| Gastrointestinal tolerability | 5–7 days | Acute malaria symptoms and blood-stage treatment can themselves cause gastrointestinal disturbances similar to those resulting from primaquine. We therefore selected days 5–7, when primaquine administration was ongoing but schizontocidal treatment had been completed and acute malaria symptoms had largely resolved. This window was chosen to better attribute gastrointestinal symptoms to primaquine exposure. |
| Haematological safety | 1–14 days | The highest risk of primaquine-induced haemolysis occurs during the first few days after treatment initiation while the drug is being administered. The first two or three days were expected to capture the most clinically relevant early haemoglobin declines. The 1–14 day window was selected to encompass the full duration of treatment, as regimens varied between 7 and 14 days. |

#### Data analysis: anti-relapse efficacy

Variables violating proportional hazards assumptions were adjusted accordingly. Patients were censored at recurrence, new antimalarial treatment, loss to follow-up, study end, outcome assessment, or a smear gap >67 days.

To detect residual confounding, we used negative control methods,^30^ using *P. falciparum* recurrence and patient body weight (see **Figure S*2*** for the rationale). Sensitivity analyses restricted the event of interest to symptomatic *P. vivax* recurrent parasitaemia and excluded unsupervised regimens, as well as data from the sole cluster-randomised trial. In studies with ≥180 days follow-up that tracked patients through multiple episodes of *P. vivax* parasitaemia, the incidence of recurrence was also analysed using Poisson regression. Rates were calculated as episodes per person-year, with censoring consistent with the Cox model.

#### Data analysis: haematological safety

The analysis of absolute fall in haemoglobin level was repeated for endpoints on days 2–3 and 5–7. Among patients with normal G6PD activity and baseline haemoglobin ≥11 g/dL, risk of anaemia by days 2–3 was modelled with Poisson regression using the same covariates, excluding G6PD level. Haematological recovery in this G6PD group was modelled with a multilevel linear model using time, daily primaquine dose, baseline haemoglobin, age, sex, log-transformed baseline parasite density, study site, and patient-specific intercepts. Splines and interaction terms were used for time and dose. The effect of daily primaquine dose on the day 7 methaemoglobin concentration and the risk of clinical methaemoglobinaemia (i.e., 10% or above) between days 1 and 14 was modelled using multivariable linear and Poisson regression, respectively; including age (years), sex (male or female), baseline parasite density (parasites per μL of blood; natural log transformed), and study site as covariates. We fitted this model by incorporating a nonlinear effect using a restricted cubic spline.

**Table S1.** PRISMA-IPD checklist.

| PRISMA-IPD<br>Section/topic | Item<br>No | Checklist item | Reported on page |
| --- | --- | --- | --- |
| Title |  |  |  |
| Title | 1 | Identify the report as a systematic review and meta-analysis of individual participant data. | 1 |
| Abstract |  |  |  |
| Structured<br>summary | 2 | Provide a structured summary including as applicable: | 2 |
|  |  | Background: state research question and main objectives, with information on participants, interventions, comparators and outcomes. |  |
|  |  | Methods: report eligibility criteria; data sources including dates of last bibliographic search or elicitation, noting that IPD were sought; methods of assessing risk of bias. |  |
|  |  | Results: provide number and type of studies and participants identified and number (%) obtained; summary effect estimates for main outcomes (benefits and harms) with confidence intervals and measures of statistical heterogeneity. Describe the direction and size of summary effects in terms meaningful to those who would put findings into practice. |  |
|  |  | Discussion: state main strengths and limitations of the evidence, general interpretation of the results and any important implications. |  |
|  |  | Other: report primary funding source, registration number and registry name for the systematic review and IPD meta-analysis. |  |
| Introduction |  |  |  |
| Rationale | 3 | Describe the rationale for the review in the context of what is already known. | 4 |
| Objectives | 4 | Provide an explicit statement of the questions being addressed with reference, as applicable, to participants, interventions, comparisons, outcomes and study design (PICOS). Include any hypotheses that relate to particular types of participant-level subgroups. | 4 |
| Methods |  |  |  |
| Protocol and registration | 5 | Indicate if a protocol exists and where it can be accessed. If available, provide registration information including registration number and registry name. Provide publication details, if applicable. | 5 |
| Eligibility criteria | 6 | Specify inclusion and exclusion criteria including those relating to participants, interventions, comparisons, outcomes, study design and characteristics (e.g. years when conducted, required minimum follow-up). Note whether these were applied at the study or individual level i.e. whether eligible participants were included (and ineligible participants excluded) from a study that included a wider population than specified by the review inclusion criteria. The rationale for criteria should be stated. | 5 |
| Identifying studies - information sources | 7 | Describe all methods of identifying published and unpublished studies including, as applicable: which bibliographic databases were searched with dates of coverage; details of any hand searching including of conference proceedings; use of study registers and agency or company databases; contact with the original research team and experts in the field; open adverts and surveys. Give the date of last search or elicitation. | 5 |
| Identifying studies - search | 8 | Present the full electronic search strategy for at least one database, including any limits used, such that it could be repeated. | 5 |
| Study selection processes | 9 | State the process for determining which studies were eligible for inclusion. | 5 |
| Data collection processes | 10 | Describe how IPD were requested, collected and managed, including any processes for querying and confirming data with investigators. If IPD were not sought from any eligible study, the reason for this should be stated (for each such study). | 5 |
|  |  | If applicable, describe how any studies for which IPD were not available were dealt with. This should include whether, how and what aggregate data were sought or extracted from study reports and publications (such as extracting data independently in duplicate) and any processes for obtaining and confirming these data with investigators. |  |
| Data items | 11 | Describe how the information and variables to be collected were chosen. List and define all study level and participant level data that were sought, including baseline and follow-up information. If applicable, describe methods of standardising or translating variables within the IPD datasets to ensure common scales or measurements across studies. | 5 |
| IPD integrity | A1 | Describe what aspects of IPD were subject to data checking (such as sequence generation, data consistency and completeness, baseline imbalance) and how this was done. | 5 |
| Risk of bias assessment in individual studies. | 12 | Describe methods used to assess risk of bias in the individual studies and whether this was applied separately for each outcome. If applicable, describe how findings of IPD checking were used to inform the assessment. Report if and how risk of bias assessment was used in any data synthesis. | 7 |
| Specification of outcomes and effect measures | 13 | State all treatment comparisons of interests. State all outcomes addressed and define them in detail. State whether they were pre-specified for the review and, if applicable, whether they were primary/main or secondary/additional outcomes. Give the principal measures of effect (such as risk ratio, hazard ratio, difference in means) used for each outcome. | 5–6 |
| Synthesis methods | 14 | Describe the meta-analysis methods used to synthesise IPD. Specify any statistical methods and models used. Issues should include (but are not restricted to):<br>• Use of a one-stage or two-stage approach.<br>• How effect estimates were generated separately within each study and combined across studies (where applicable).<br>• Specification of one-stage models (where applicable) including how clustering of patients within studies was accounted for.<br>• Use of fixed or random effects models and any other model assumptions, such as proportional hazards.<br>• How (summary) survival curves were generated (where applicable).<br>• Methods for quantifying statistical heterogeneity (such as $I^2$ and $t^2$).<br>• How studies providing IPD and not providing IPD were analysed together (where applicable).<br>• How missing data within the IPD were dealt with (where applicable). | 5–6 |
| Exploration of variation in effects | A2 | If applicable, describe any methods used to explore variation in effects by study or participant level characteristics (such as estimation of interactions between effect and covariates). State all participant-level characteristics that were analysed as potential effect modifiers, and whether these were pre-specified. | 5–6 |
| Risk of bias across studies | 15 | Specify any assessment of risk of bias relating to the accumulated body of evidence, including any pertaining to not obtaining IPD for particular studies, outcomes or other variables. | 8 |
| Additional analyses | 16 | Describe methods of any additional analyses, including sensitivity analyses. State which of these were pre-specified. | 5–6 |
| **Results** |  |  |  |
| Study selection and IPD obtained | 17 | Give numbers of studies screened, assessed for eligibility, and included in the systematic review with reasons for exclusions at each stage. Indicate the number of studies and participants for which IPD were sought and for which IPD were obtained. For those studies where IPD were not available, give the numbers of studies and participants for which aggregate data were available. Report reasons for non-availability of IPD. Include a flow diagram. | 8 |
| Study characteristics | 18 | For each study, present information on key study and participant characteristics (such as description of interventions, numbers of participants, demographic data, unavailability of outcomes, funding source, and if applicable duration of follow-up). Provide (main) citations for each study. Where applicable, also report similar study characteristics for any studies not providing IPD. | 8 |
| IPD integrity | A3 | Report any important issues identified in checking IPD or state that there were none. | 8 |
| Risk of bias within studies | 19 | Present data on risk of bias assessments. If applicable, describe whether data checking led to the up-weighting or down-weighting of these assessments. Consider how any potential bias impacts on the robustness of meta-analysis conclusions. | 8 |
| Results of individual studies | 20 | For each comparison and for each main outcome (benefit or harm), for each individual study report the number of eligible participants for which data were obtained and show simple summary data for each intervention group (including, where applicable, the number of events), effect estimates and confidence intervals. These may be tabulated or included on a forest plot. | 8 |
| Results of syntheses | 21 | Present summary effects for each meta-analysis undertaken, including confidence intervals and measures of statistical heterogeneity. State whether the analysis was pre-specified, and report the numbers of studies and participants and, where applicable, the number of events on which it is based. | 8–9 |
|  |  | When exploring variation in effects due to patient or study characteristics, present summary interaction estimates for each characteristic examined, including confidence intervals and measures of statistical heterogeneity. State whether the analysis was pre-specified. State whether any interaction is consistent across trials. |  |
|  |  | Provide a description of the direction and size of effect in terms meaningful to those who would put findings into practice. |  |
| Risk of bias across studies | 22 | Present results of any assessment of risk of bias relating to the accumulated body of evidence, including any pertaining to the availability and representativeness of available studies, outcomes or other variables. | 8 |
| Additional analyses | 23 | Give results of any additional analyses (e.g. sensitivity analyses). If applicable, this should also include any analyses that incorporate aggregate data for studies that do not have IPD. If applicable, summarise the main meta-analysis results following the inclusion or exclusion of studies for which IPD were not available. | 8–9 |
| **Discussion** |  |  |  |
| Summary of evidence | 24 | Summarise the main findings, including the strength of evidence for each main outcome. | 10 |
| Strengths and limitations | 25 | Discuss any important strengths and limitations of the evidence including the benefits of access to IPD and any limitations arising from IPD that were not available. | 10–12 |
| Conclusions | 26 | Provide a general interpretation of the findings in the context of other evidence. | 10–12 |
| Implications | A4 | Consider relevance to key groups (such as policy makers, service providers and service users). Consider implications for future research. | 10–12 |

| Funding |  |  |  |
| --- | --- | --- | --- |
| Funding | 27 | Describe sources of funding and other support (such as supply of IPD), and the role in the systematic review of those providing such support. | 7 |

**Table S2.**
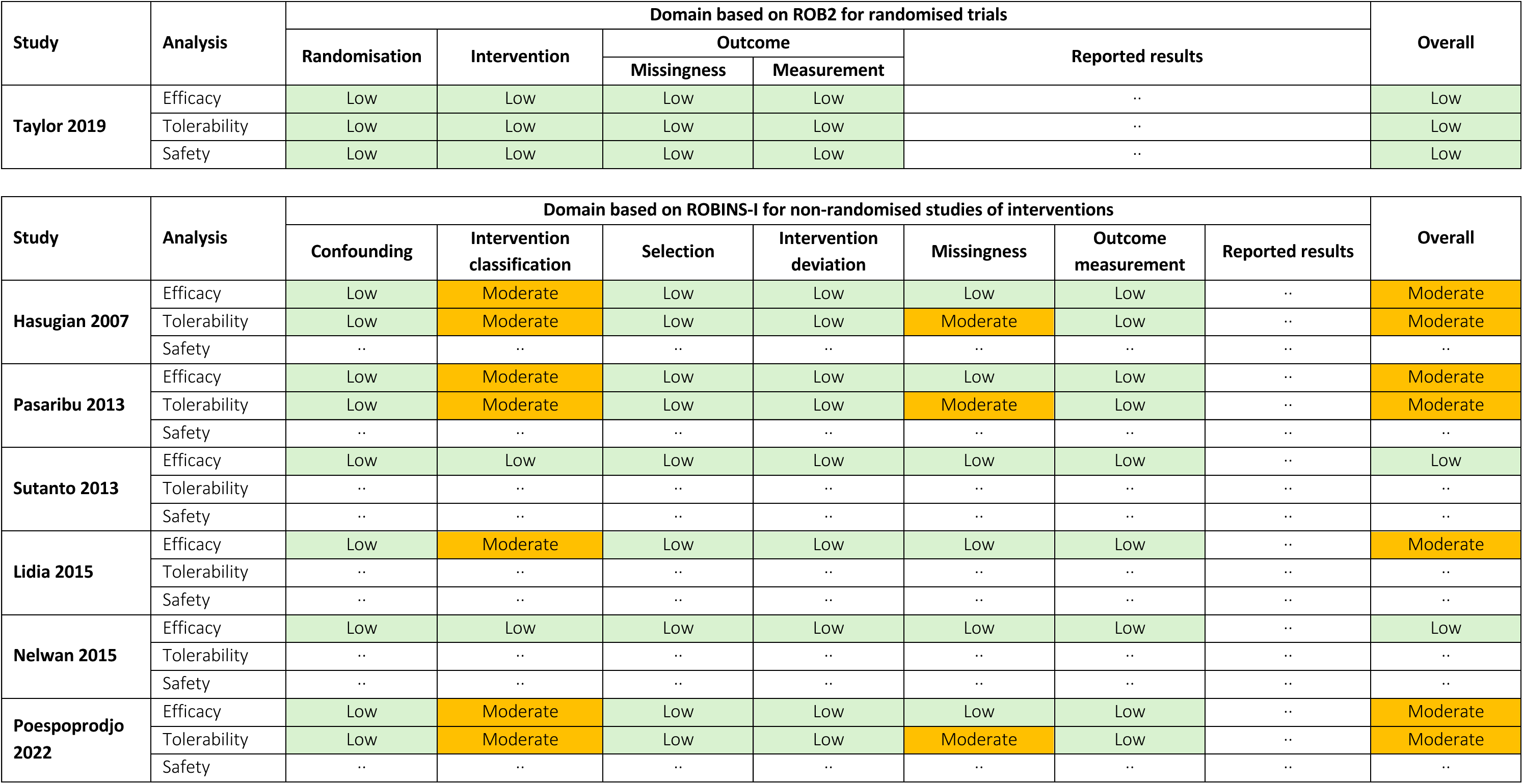
Risk of bias assessment.

**Figure S1.**
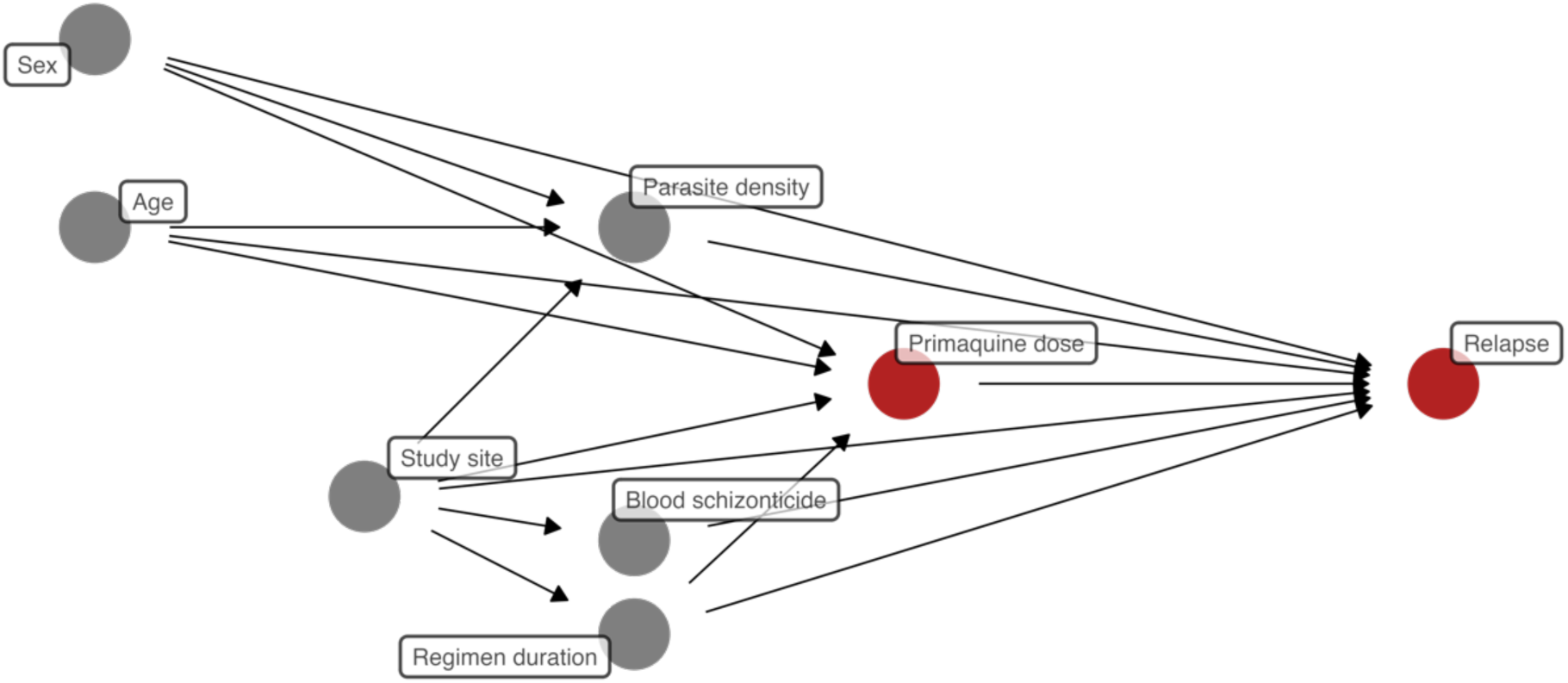
Causal directed acyclic graph showing the relationship between the two factors of interest (red nodes): primaquine mg/kg total dose and the risk of *P. vivax* relapse. Under this causal framework, age, sex, and study site are considered potential confounders of the dose–relapse association. These variables may act as proxies for underlying host factors (e.g., body weight, acquired immunity, treatment-seeking behaviour, CYP2D6 polymorphisms) and geographical or parasite-related characteristics (e.g., strain relapse periodicity, transmission intensity, blood-stage drug resistance) that influence both primaquine dosing and relapse risk. Parasite density, blood schizontocidal treatment, and regimen duration are included as factors that may affect relapse risk and/or influence treatment allocation. The directed acyclic graph informed selection of adjustment variables in the regression models.

**Figure S2.**
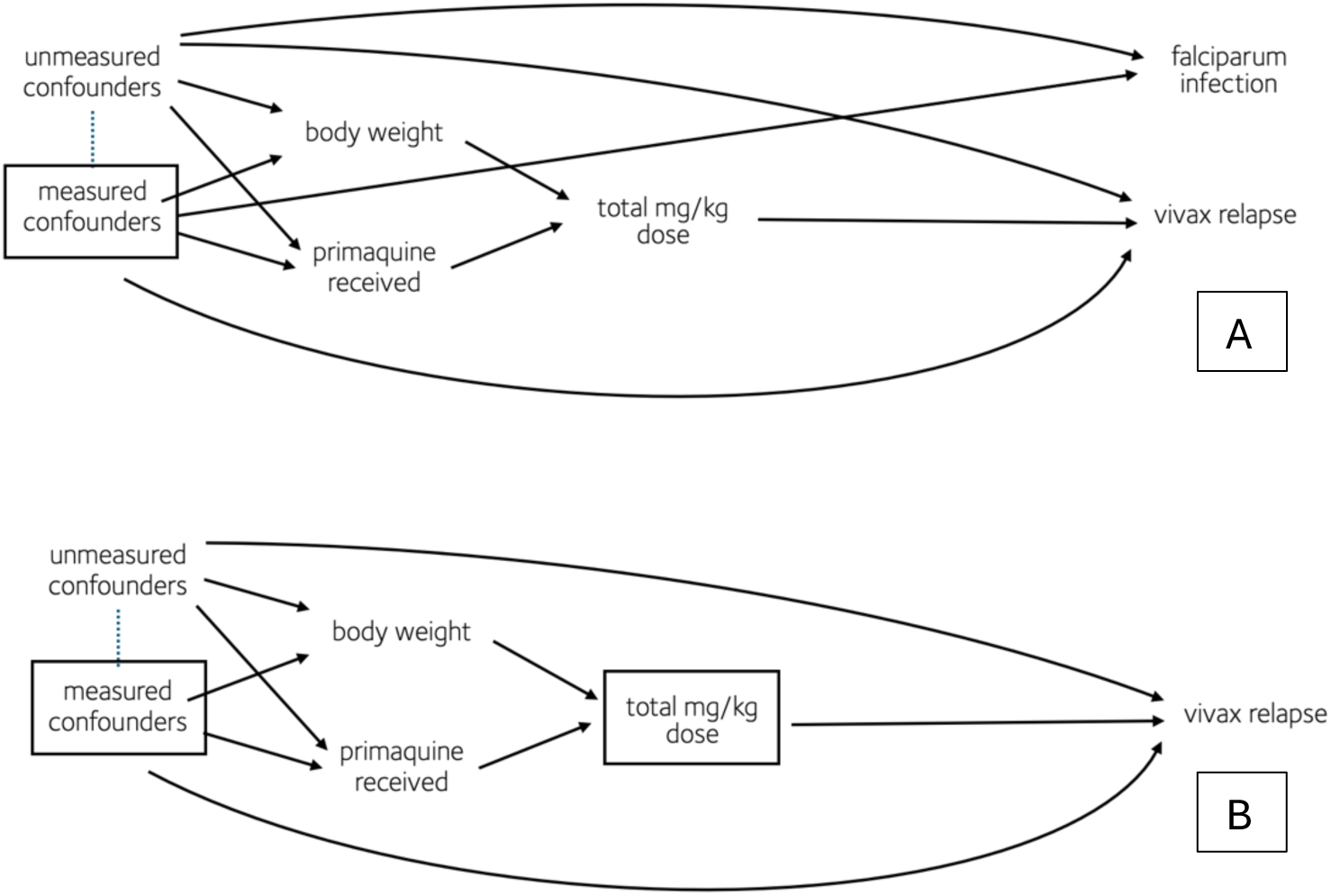
Causal directed acyclic graphs illustrating the use of *P. falciparum* infection and body weight as negative controls to detect residual confounding. A. <u>Outcome negative control</u>: The presence of *P. falciparum* re-infection in patients treated for *P. vivax* mono-infection during follow-up in areas with local transmission should be independent of the total primaquine dose received, provided that measured confounding is adequately adjusted for. However, if substantial residual confounding is not sufficiently controlled (either due to inadequate adjustment of measured confounding or the presence of unmeasured confounding), a spurious causal effect of primaquine dose on the risk of *P. falciparum* infection may become apparent. B. <u>Exposure negative control</u>: Among patients with *P. vivax* who received no primaquine (thereby excluding the essential anti-hypnozoite component of treatment) the risk of *P. vivax* relapse should be independent of the total primaquine dose received, provided that measured confounding is adequately adjusted for. However, if substantial residual confounding is not sufficiently controlled (either due to inadequate adjustment of measured confounding or the presence of unmeasured confounding) a spurious causal effect of body weight on the risk of *P. vivax* relapse may become apparent. A node within a square represents an adjustment, either through model stratification or restriction in our analysis. A dotted line indicates that two nodes may be related due to some common causes.

**Figure S3.**
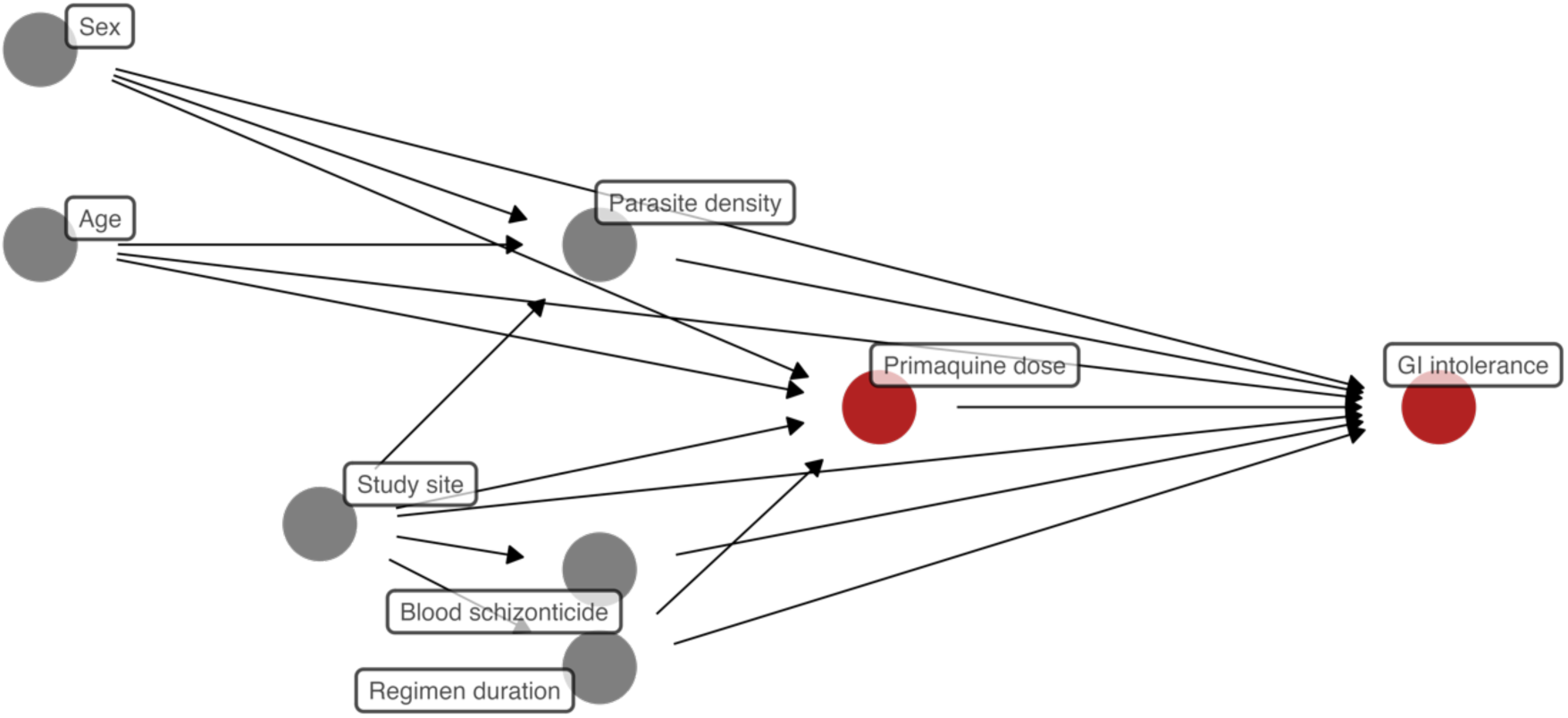
Causal directed acyclic graph showing the relationship between the two factors of interest (red nodes): primaquine mg/kg daily dose and the risk of gastrointestinal (GI) discomfort. Under this causal framework, age, sex, and study site are considered potential confounders of the dose–GI causal association. These variables may act as proxies for underlying host characteristics (e.g., body weight, treatment adherence, CYP2D6 polymorphisms) and contextual or geographical factors that influence both dosing and susceptibility to gastrointestinal symptoms. The directed acyclic graph informed covariate selection for adjustment in the tolerability analyses.

**Figure S4.**
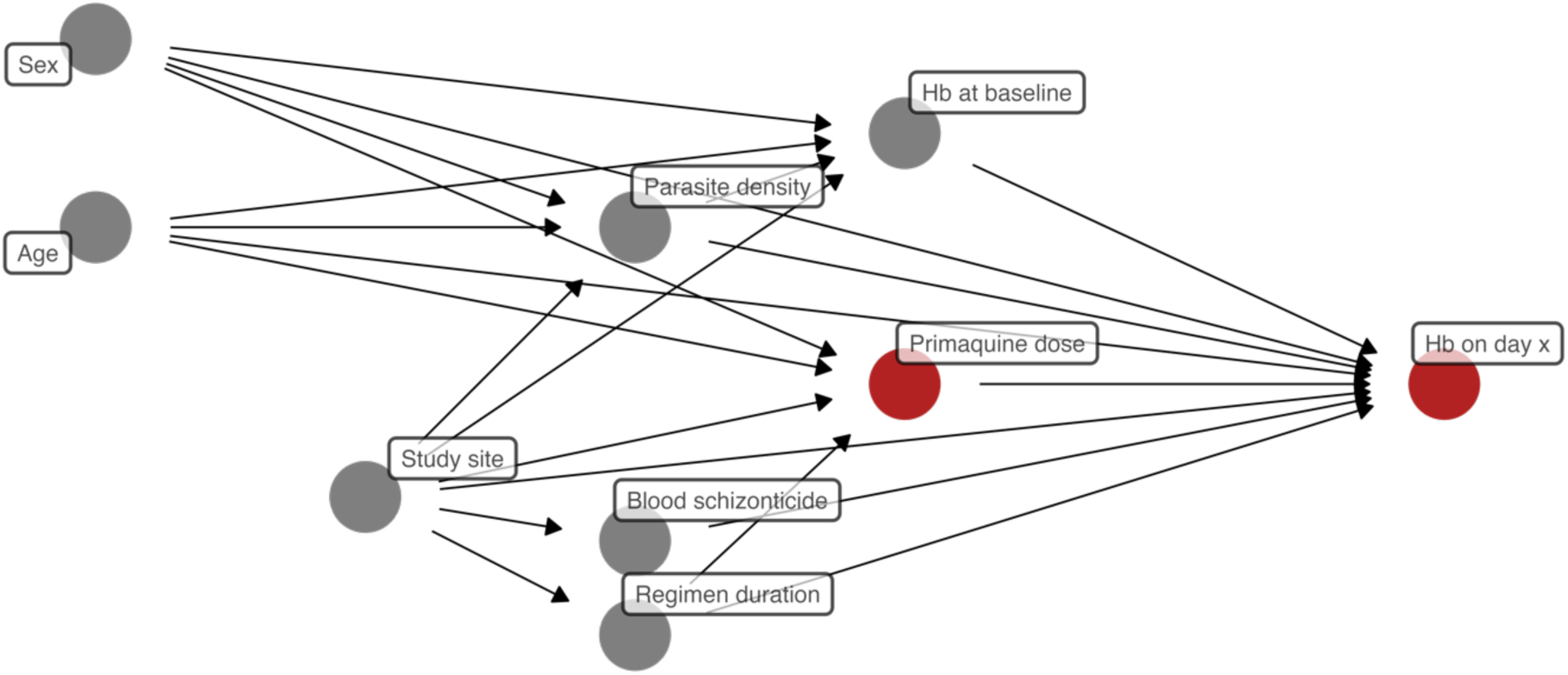
Causal directed acyclic graph showing the relationship between the two factors of interest (red nodes): primaquine mg/kg daily dose and haemolysis. Under this causal framework, age, sex, and study site are considered potential confounders of the dose–haemolysis association. These variables may act as proxies for underlying host factors (e.g., body weight, baseline haemoglobin, G6PD status, treatment behaviours, CYP2D6 polymorphisms) and geographical or epidemiological characteristics that influence both dosing and susceptibility to haemolysis. The directed acyclic graph informed covariate selection for adjustment in the haematological safety analyses.

**Figure S5.**
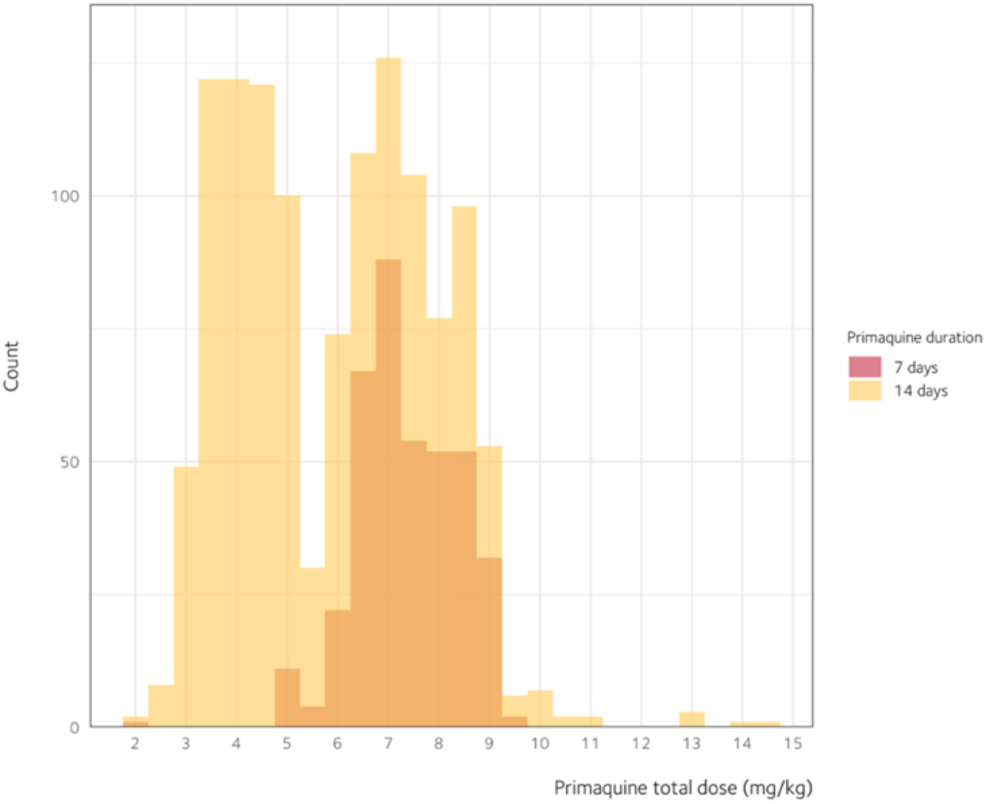
Distribution of primaquine total dose by primaquine duration. In the 14-day primaquine regimen, the observed two peaks reflect the targeted total primaquine dose of 3·5 and 7 mg base per kg body weight.

**Figure S6.**
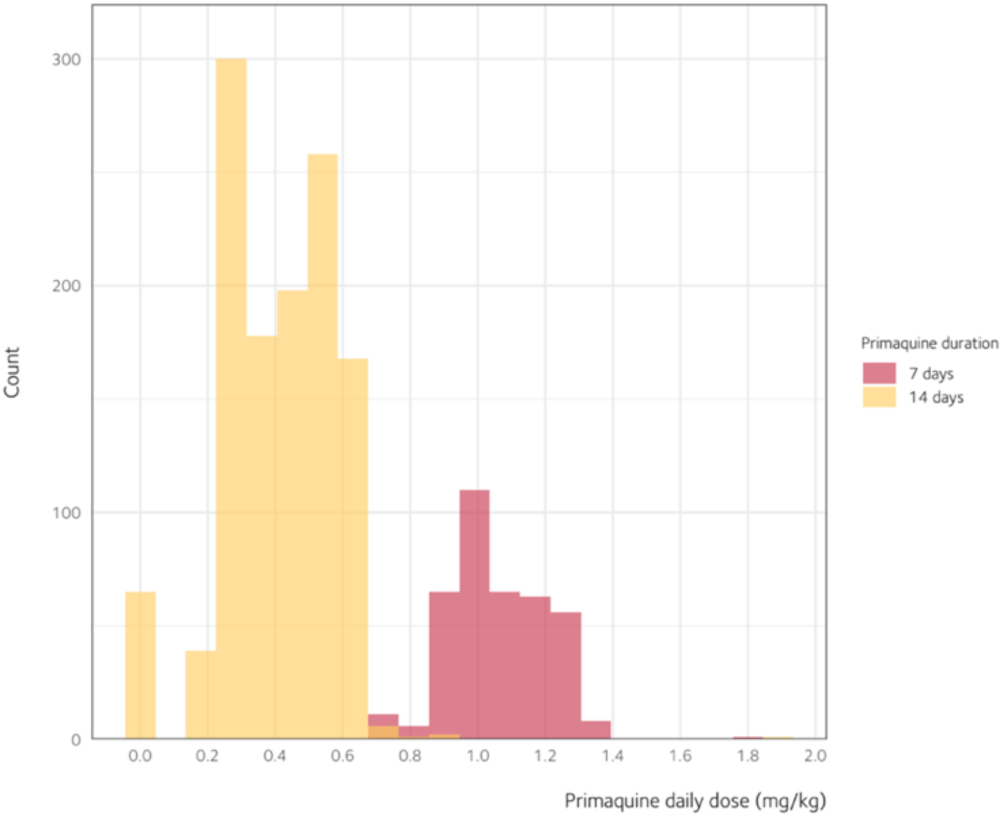
Distribution of primaquine daily dose by primaquine regimen.

**Table S3.**
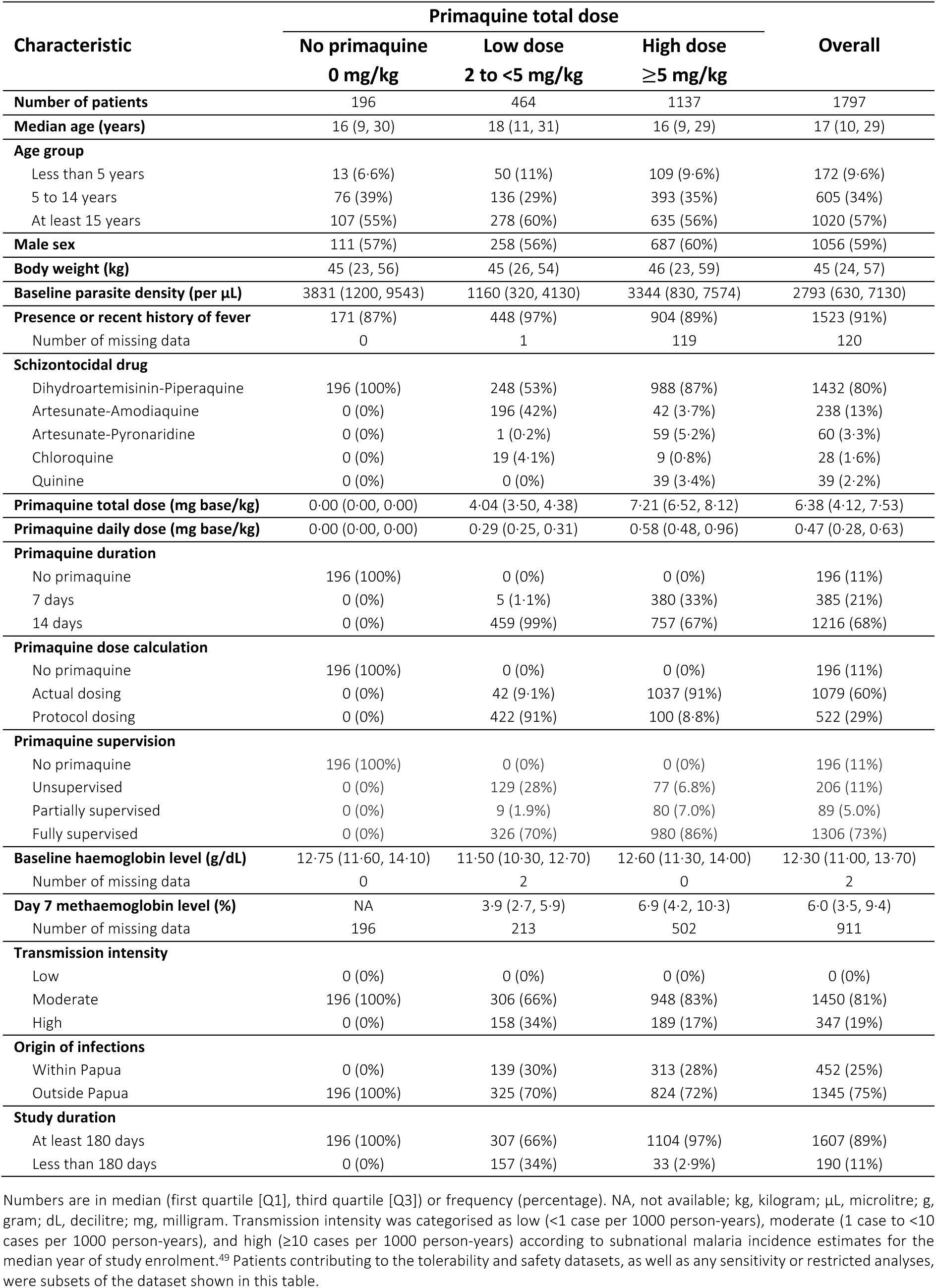
Patient characteristics by primaquine total dose group.

**Table S4.** Studies included in analysis.

| Paper | Study site | Latitude | Longitude | Year start | Year end | MAP incidence rate (per 1000 person-years) | Transmission intensity# | Relapse periodicity§ |
| --- | --- | --- | --- | --- | --- | --- | --- | --- |
| **Hasugian 2007<sup>24</sup>** | Timika | −4.61 | 136.85 | 2005 | 2005 | 22.61 | High | High |
| **Pasaribu 2013<sup>14</sup>** | Tanjung Leidong | 2.77 | 99.98 | 2011 | 2011 | 2.75 | Moderate | High |
| **Sutanto 2013<sup>13</sup>** | Lumajang | −8.13 | 113.22 | 2010 | 2011 | 36.88* | High | High |
| **Lidia 2015<sup>25</sup>** | Kupang | −10.18 | 123.61 | 2013 | 2013 | 15.22 | High | High |
| **Nelwan 2015<sup>11</sup>** | Sragen | −7.42 | 111.02 | 2013 | 2013 | 42.44* | High | High |
| **Taylor 2019<sup>15</sup>** | Hanura | −5.53 | 105.24 | 2015 | 2017 | 1.01 | Moderate | High |
| **Taylor 2019<sup>15</sup>** | Tanjung Leidong | 2.77 | 99.98 | 2015 | 2017 | 1.03 | Moderate | High |
| **Poespoprodjo 2022<sup>12</sup>** | Timika | −4.45 | 136.98 | 2015 | 2018 | 56.43 | High | High |

MAP malaria atlas project. § Relapse periodicity was categorised as high (median relapse periodicity of 47 days or less) and low (median relapse periodicity of more than 47);<sup>52</sup> # Transmission intensity was categorised as low (<1 case per 1000 person-years), moderate (1 case to <10 cases per 1000 person-years), and high (≥10 cases per 1000 person-years) according to subnational malaria incidence estimates for the median year of study enrolment.<sup>49</sup> \* Based on the location where patients were infected by *P. vivax* in Papua.

**Table S5.** Studies that were eligible for analysis but not included for data pooling.

| Characteristic | Study |  |  |
| --- | --- | --- | --- |
|  | Maguire et al.<sup>27*</sup> | Arcelia et al.<sup>28</sup> | Sutanto et al.<sup>26</sup> |
| Year published | 2006 | 2023 | 2023 |
| Number of treatment arms | 2 | 1 | 3 |
| Region origin of infection | Papua | Outside Papua | Papua |
| Number of sites | 1 | 1 | 2 |
| Follow-up (days) | 28 | 28 | 180 |
| Randomised | Yes | No | Yes |
| Recruitment period | 1996–1999 | 2019–2020 | 2018–2019 |
| Treatment arms | (1) Chloroquine + 14-day, low dose primaquine<br>(2) Mefloquine + 14-day, low dose primaquine | (1) Dihydroartemisinin-Piperaquine + 14-day, low dose primaquine | (1) Dihydroartemisinin-Piperaquine<br>(2) Dihydroartemisinin-Piperaquine + 14-day, low dose primaquine<br>(3) Dihydroartemisinin-Piperaquine + Tafenoquine |
| *P. vivax* patients enrolled | 125* | 60 | 150 |
| Treated with primaquine | 125* | 60 | 50 |
| Primaquine supervision | NA | NA | Yes |
| Sex (primaquine receiving arm) | 68% Male* | NA | 100% Male |
| Age (in years, primaquine receiving arm) | Mean = 22·3* | 30 Children (2 to 18 years)<br>30 Adults | Mean = 28·6 |
| Reason for exclusion | Missing minimum data | Data not available by 28 February 2025 | Data not available by 28 February 2025 |

\* Based on contributed IPD that may include both randomised trials and prospective cohorts not part of the trial itself. NA, not available; IPD, individual patient data.

**Table S6.**
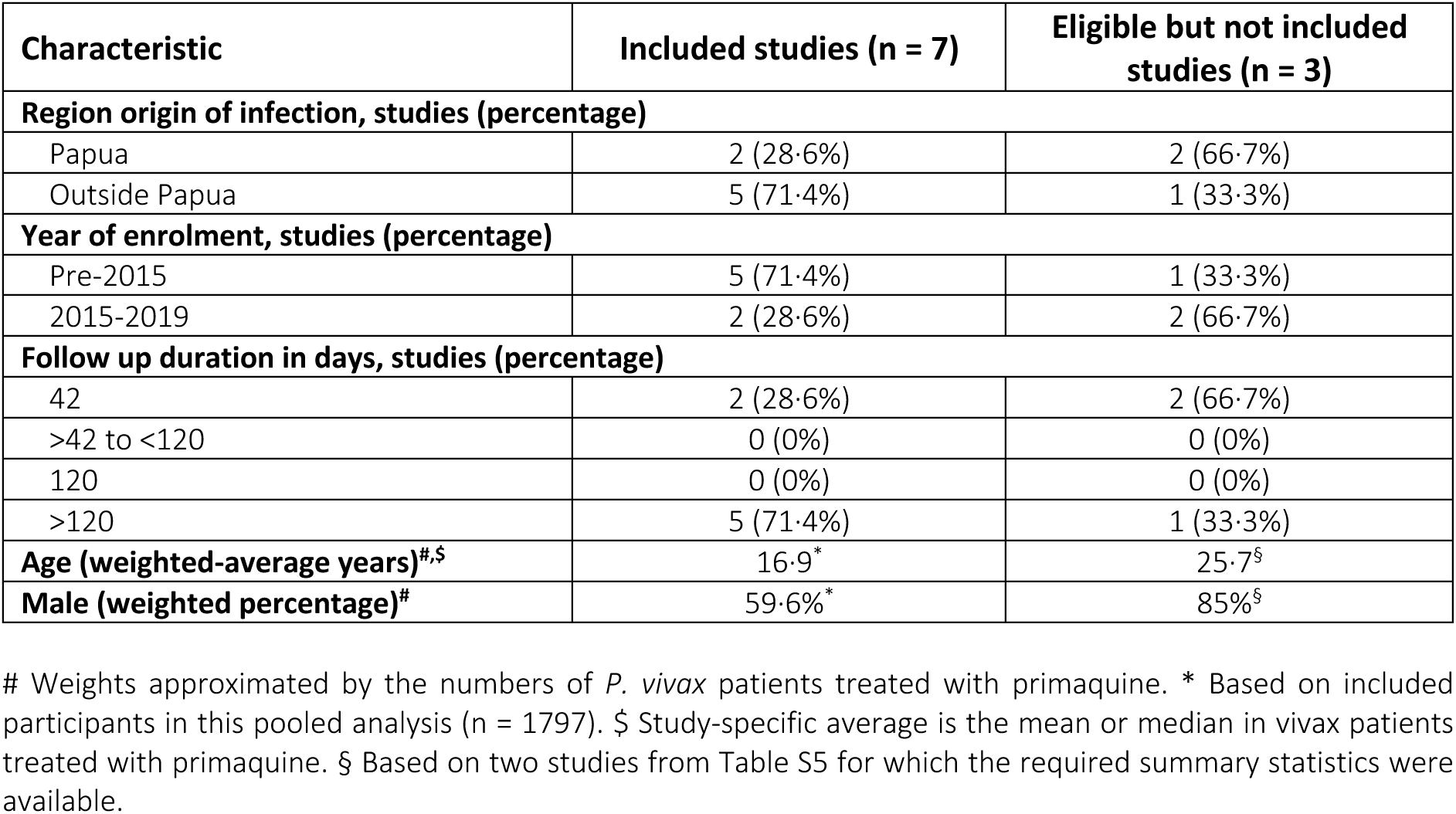
Comparison of patient characteristics who received primaquine between included studies and eligible but not available studies.

| Characteristic | Included studies (n = 7) | Eligible but not included studies (n = 3) |
| --- | --- | --- |
| **Region origin of infection, studies (percentage)** |  |  |
| Papua | 2 (28.6%) | 2 (66.7%) |
| Outside Papua | 5 (71.4%) | 1 (33.3%) |
| **Year of enrolment, studies (percentage)** |  |  |
| Pre-2015 | 5 (71.4%) | 1 (33.3%) |
| 2015-2019 | 2 (28.6%) | 2 (66.7%) |
| **Follow up duration in days, studies (percentage)** |  |  |
| 42 | 2 (28.6%) | 2 (66.7%) |
| >42 to <120 | 0 (0%) | 0 (0%) |
| 120 | 0 (0%) | 0 (0%) |
| >120 | 5 (71.4%) | 1 (33.3%) |
| **Age (weighted-average years)<sup>#,§</sup>** | 16.9<sup>*</sup> | 25.7<sup>§</sup> |
| **Male (weighted percentage)<sup>#</sup>** | 59.6%<sup>*</sup> | 85%<sup>§</sup> |

**Figure S7.**
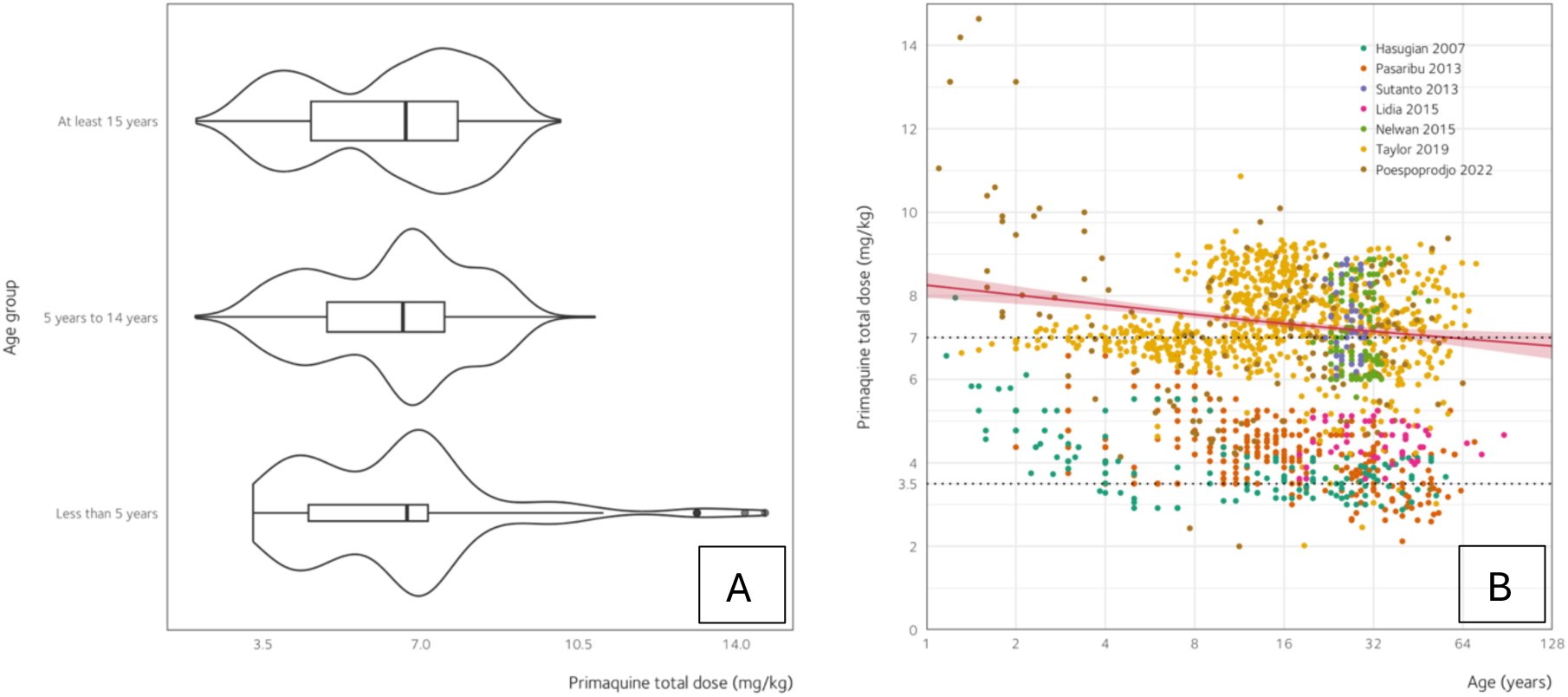
Primaquine total dose received across different patient age. A. While the median average across the three age groups was similar, patients under five years old were more likely to receive a higher total dose (relative to a target dose) than older patients, as also depicted in panel B. In B, each dot represents an individual patient. The red curve denotes a fitted regression line between age and primaquine total dose. Dotted line represents the target dose of 3·5 or 7 mg base per kg body weight. This may reflect the relatively greater difficulty of administering primaquine in very young patients. For panel B, the horizontal axis is shown on a logarithmic scale.

**Figure S8.**
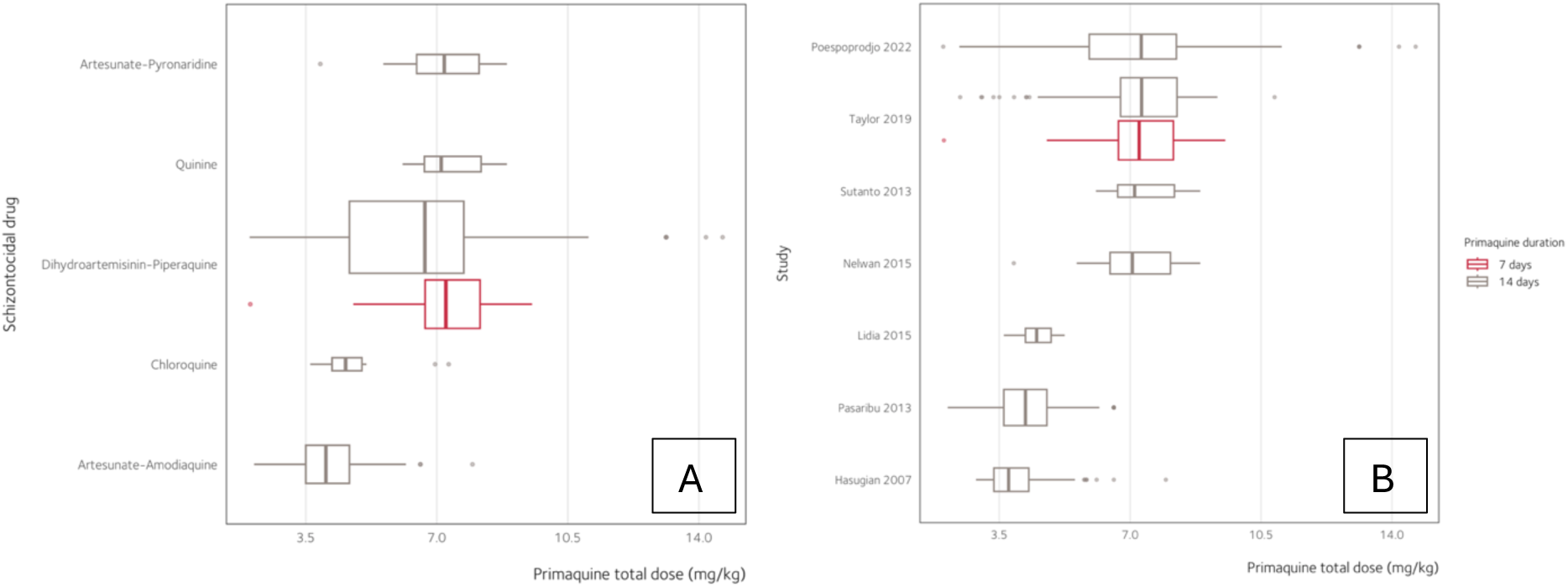
Primaquine total dose by schizontocidal drug (A) and study (B)

**Figure S9.**
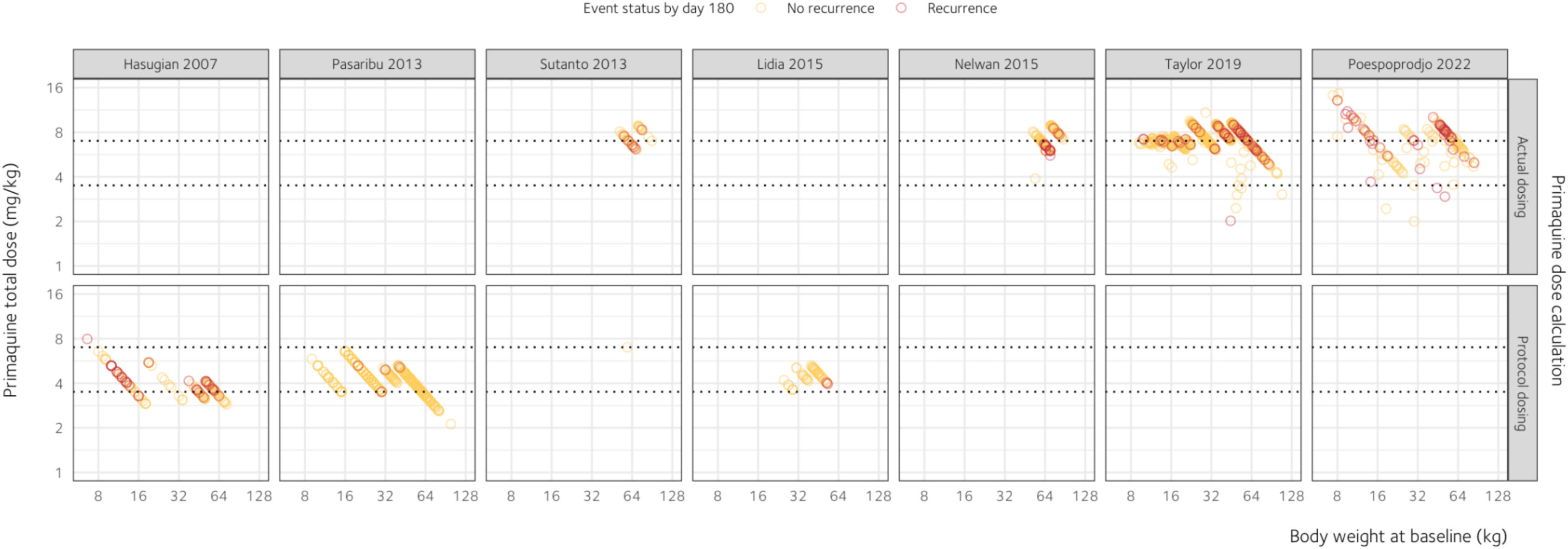
Primaquine total dose by body weight in patients receiving primaquine. Dotted line represents the total dose targets of 3·5 or 7 mg/kg. For Hasugian 2007 and Lidia 2015, the follow up duration was less than 180 days. Event status indicates whether the patient developed a *P. vivax* recurrence. Horizontal and vertical axes are shown on a logarithmic scale.

**Figure S10.**
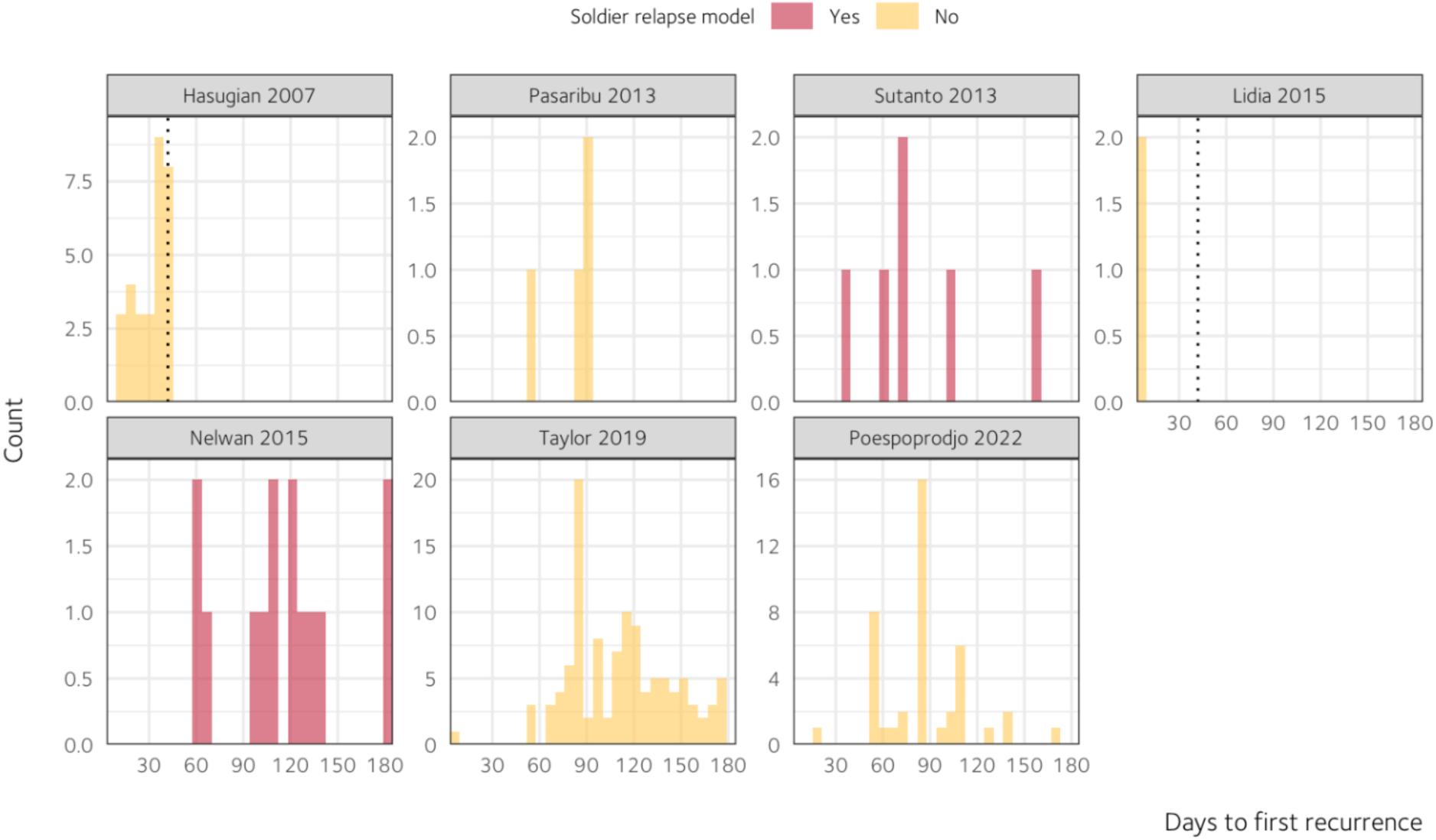
Distributions of the days to first *P. vivax* recurrence by study. For Hasugian 2007 and Lidia 2015, the dotted line represents the end of active follow up. The other studies monitored patients for at least 180 days.

**Table S7.** Model estimates underlying the main results.

| Model | Coefficient | Standard error |
| --- | --- | --- |
| **Efficacy (Cox proportional hazards regression)** |  |  |
| Primaquine total dose (spline term 1) | −0.3185 | 0.0416 |
| Primaquine total dose (spline term 2) | 0.1794 | 0.0494 |
| Log baseline parasite density (linear term) | 0.2154 | 0.0514 |
| Age (linear term) | −0.0305 | 0.0108 |
| Age × time | 0.0003 | 0.0001 |
| **Tolerability (Poisson regression)** |  |  |
| Primaquine daily dose (spline term 1) | 1.9431 | 0.7109 |
| Primaquine daily dose (spline term 2) | −1.2393 | 0.9385 |
| Log baseline parasite density (linear term) | 0.0380 | 0.0654 |
| Age (linear term) | 0.0214 | 0.0064 |
| Sex (male vs. female) | −0.2056 | 0.2055 |
| Intercept | −4.0943 | 0.6799 |
| **Safety (linear regression)** |  |  |
| Primaquine daily dose (linear term) | −8.1201 | 6.4840 |
| Log<sub>2</sub> G6PD activity (spline term 1) | 0.5708 | 0.9312 |
| Log<sub>2</sub> G6PD activity (spline term 2) | 0.8153 | 2.7176 |
| Log<sub>2</sub> G6PD activity (spline term 3) | −8.5553 | 10.5350 |
| Baseline haemoglobin (linear term) | −0.3670 | 0.0207 |
| Age (linear term) | 0.0006 | 0.0024 |
| Sex (male vs. female) | 0.2286 | 6.9952 |
| Study site | 0.0877 | 0.0735 |
| Baseline parasite density (linear term) | 0.0086 | 0.0208 |
| Primaquine daily dose × log<sub>2</sub> G6PD activity (spline 1) | 1.3102 | 1.0217 |
| Primaquine daily dose × log<sub>2</sub> G6PD activity (spline 2) | −5.9293 | 3.2047 |
| Primaquine daily dose × log<sub>2</sub> G6PD activity (spline 3) | 27.0140 | 12.6741 |
| Primaquine daily dose × sex | 11.8332 | 8.4664 |
| log<sub>2</sub> G6PD activity (spline 1) × sex | 0.0355 | 1.1027 |
| log<sub>2</sub> G6PD activity (spline 2) × sex | −1.3898 | 3.4900 |
| log<sub>2</sub> G6PD activity (spline 3) × sex | 10.0486 | 14.1048 |
| Primaquine daily dose × log<sub>2</sub> G6PD activity (spline 1) × sex | −1.8645 | 1.3382 |
| Primaquine daily dose × log<sub>2</sub> G6PD activity (spline 2) × sex | 4.9666 | 4.4386 |
| Primaquine daily dose × log<sub>2</sub> G6PD activity (spline 3) × sex | −20.2518 | 18.0407 |
| Intercept | −0.6788 | 5.9135 |

In the efficacy model, to relax certain modelling assumptions (e.g., proportional hazards and baseline hazards across sites), study site and sex were specified as stratification variables, and an age–time interaction term was included. Coefficient estimates from the Cox and Poisson models represent log hazard ratios and log risk ratios, respectively.

**Figure S11.**
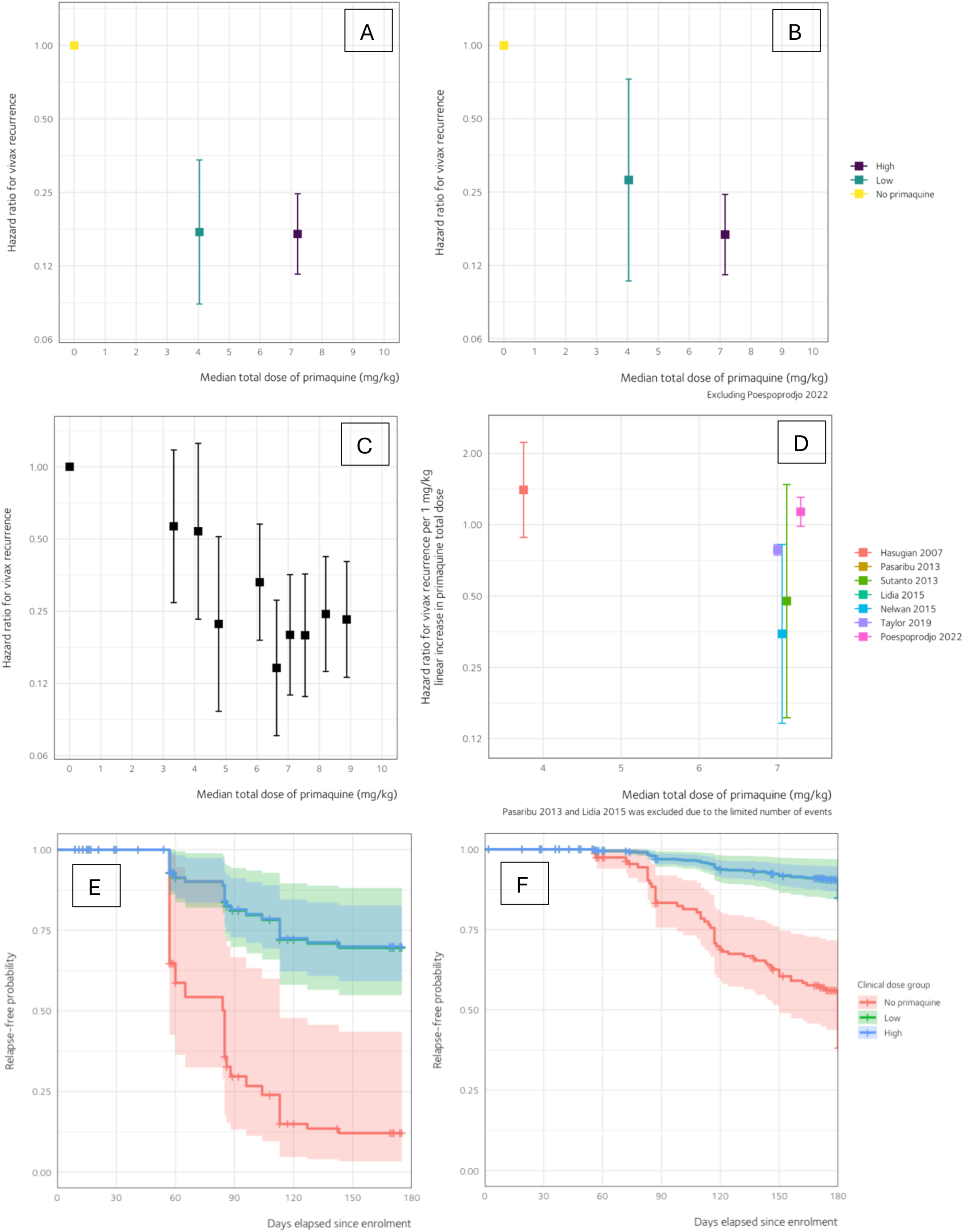
*P. vivax* recurrence at day 180 by different groups. By (A) clinical total dose group in the primary efficacy dataset, n = 1797; (B) Clinical total dose group excluding the cluster randomised trial,^12^ n = 1641; (C) Decile with 10 equal sized groups in the primary efficacy dataset, n = 1797; and (D) Excluding two studies^14,25^ due to the limited number of events, n = 1415. The reference value, at which the hazard ratio equals one, was set at 0 mg/kg. The whisker shows 95% confidence intervals. Estimates were derived from a multivariable Cox proportional hazards model. The vertical axis for A–D is shown on a logarithmic scale. Based on the model underlying panel (A), panels (E) and (F) display the model-implied relapse-free probabilities in different total dose groups, separately as examples in Papua and Non-Papua settings, respectively. To estimate these survival probabilities, continuous covariates were set at their median values, and sex was set to male. This dose–response relationship was broadly consistent across alternative dose groupings and sensitivity analyses.

**Figure S12.**
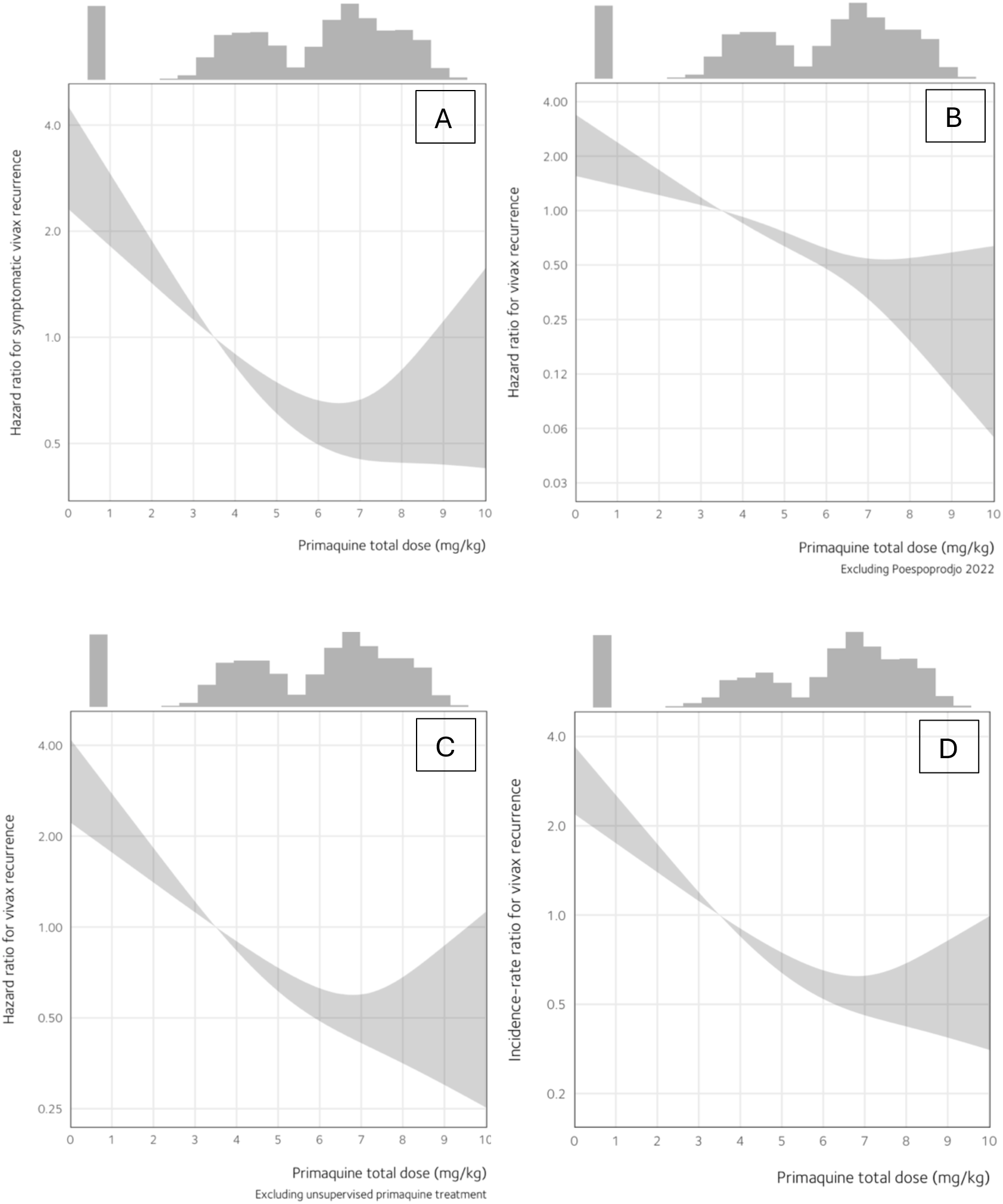
Sensitivity analyses of the hazard ratio across primaquine total dose. (A) Using symptomatic *P. vivax* recurrences as the endpoint event, n = 1794; (B) Excluding the cluster randomised trial,^12^ n = 1641; (C) Excluding unsupervised primaquine treatment, n = 1591; (D) Estimating the incidence rate ratio in patients followed for multiple *P. vivax* episodes, n = 1607. The reference value, at which the hazard ratio equals one, was set at the target low dose of 3·5 mg/kg. The shaded region shows 95% confidence intervals. Estimates were derived from a multivariable Cox proportional hazards model (A, B, C) or a multivariable Poisson model (D). A restricted cubic spline with three knots was specified on primaquine total dose to allow for a non-linear trend. The histogram along the top margin shows the distribution of primaquine daily doses in the model data, with the leftmost bar representing patients who were treated without primaquine (i.e., 0 mg/kg). The vertical axis is shown on a logarithmic scale. This dose–response relationship was broadly consistent across alternative dose groupings and sensitivity analyses.

### List S3. Negative control results

Negative control methods for exposure (AHR of *P. vivax* recurrence associated with a 1 kg increase in body weight in patients treated without primaquine = 1.1; 95% CI 0.98 to 1.03; p = 0.60) and outcome (AHR of *P. falciparum* infection associated with a 1 mg/kg increase in primaquine total dose = 1.03; 95% CI 0.94 to 1.14; p = 0.48) suggest that residual confounding in our anti-relapse effect estimates was minimal.

**Figure S13.**
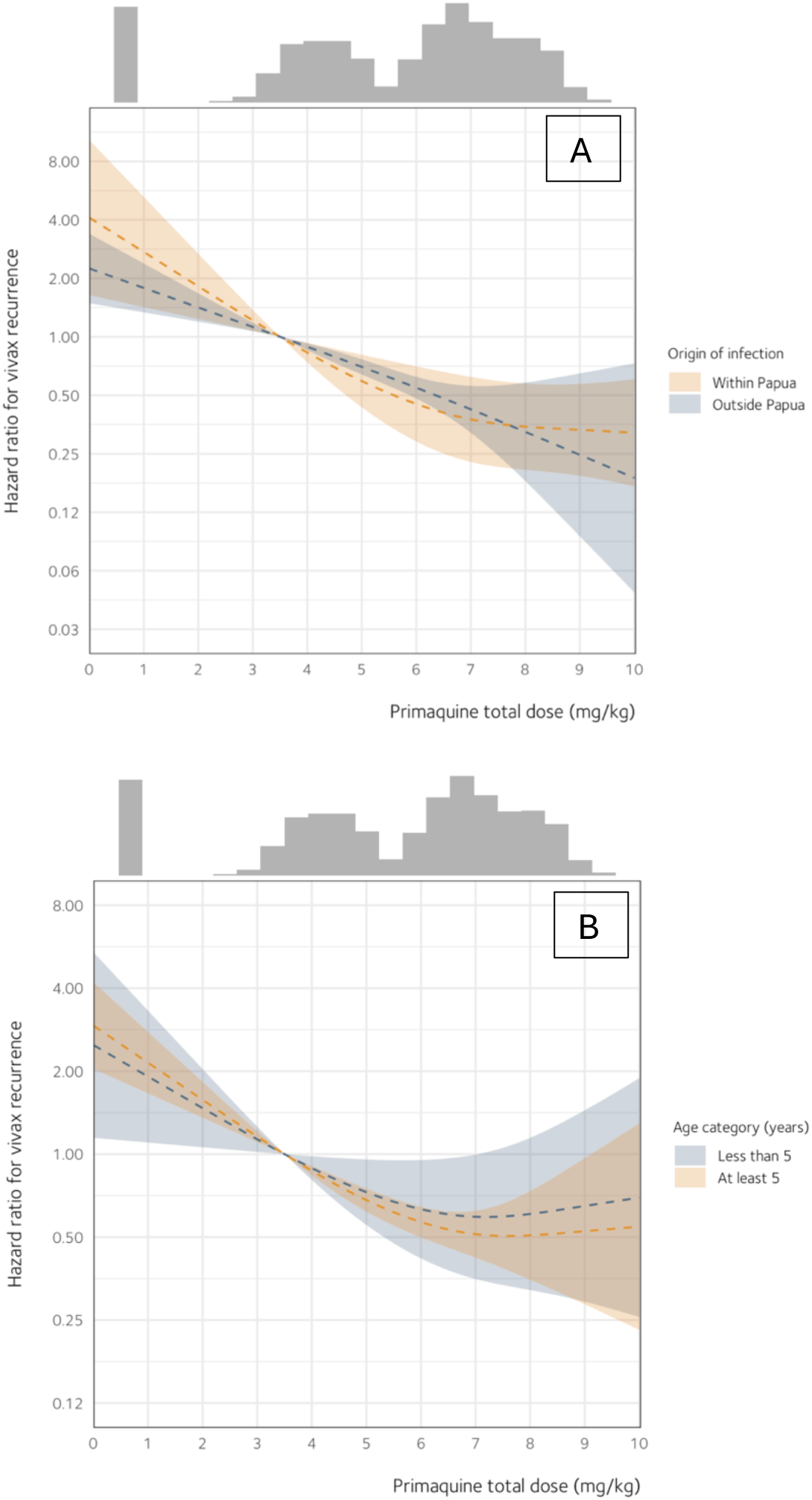
Estimated hazard ratios of *P. vivax* recurrence at day 180, assuming effect modification by (A) origin of infection and (B) age category. The reference value, at which the hazard ratio equals one, was set at the target low dose of 3·5 mg/kg. The shaded region shows 95% confidence intervals. Estimates were derived from a multivariable Cox proportional hazards model. A restricted cubic spline with three knots was specified on primaquine total dose to allow for a non-linear trend. The histogram along the top margin shows the distribution of primaquine daily doses in the model data (n = 1797), with the leftmost bar representing patients who were treated without primaquine (i.e., 0 mg/kg). The vertical axis is shown on a logarithmic scale.

**Figure S14.**
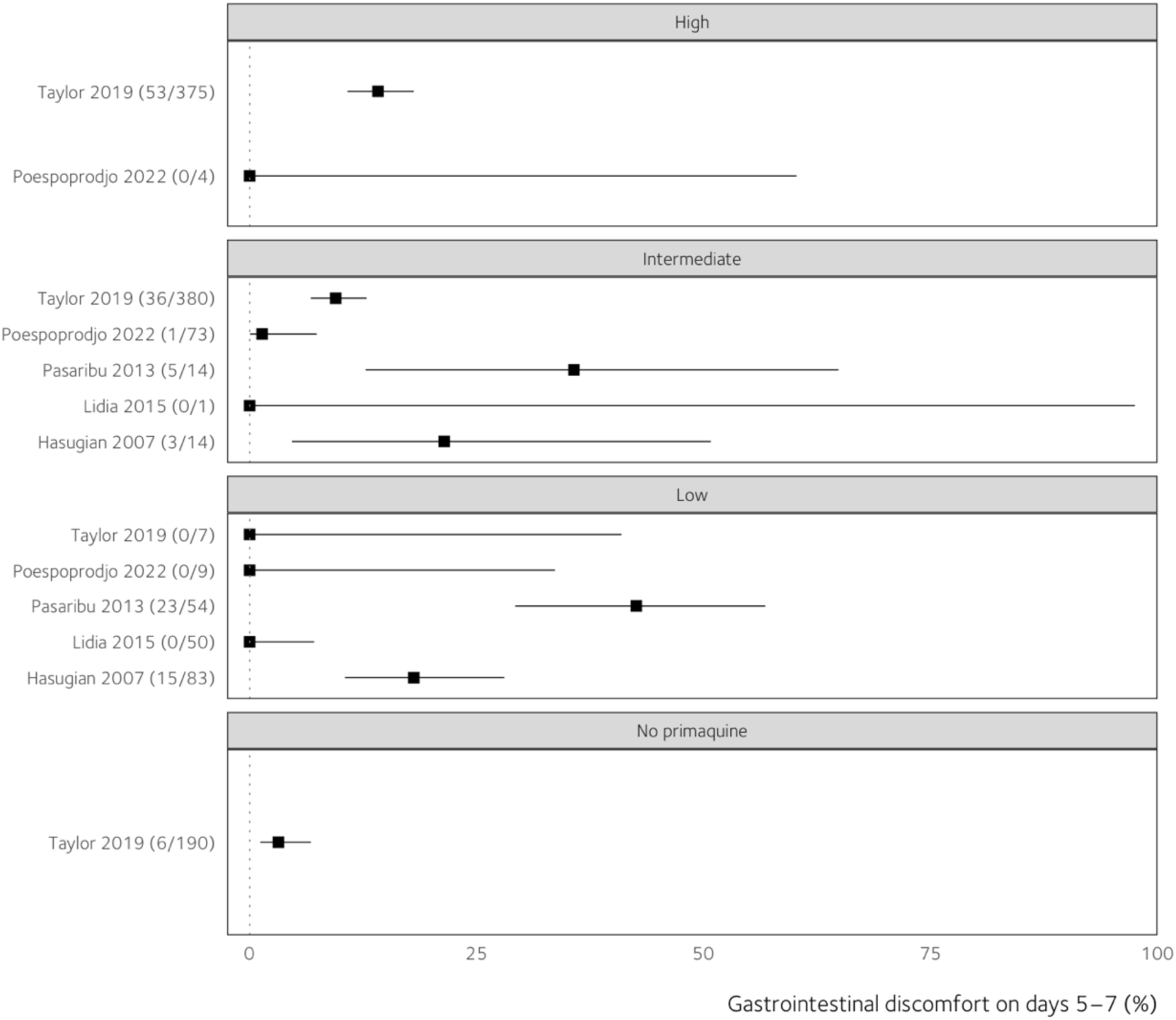
Heterogeneity in percentages of gastrointestinal discomfort on days 5–7 by study and primaquine daily dose. This variability may stem from the relatively subjective nature of assessing this endpoint. The solid square and horizontal line represent the point and 95% confidence interval estimates.

**Table S8.** Pooled counts and percentages of patients who experienced gastrointestinal discomfort.

| Primaquine daily dose | Days of observation |  |  |
| --- | --- | --- | --- |
|  | Baseline | 1–2 | 5–7 |
| No primaquine | 0/0 (…) | 0/0 (…) | 6/190 (3.2%) |
| Low | 9/335 (2.7%) | 46/335 (13.7%) | 38/203 (18.7%) |
| Intermediate | 309/584 (52.9%) | 192/583 (32.9%) | 45/482 (9.3%) |
| High | 298/376 (79.3%) | 189/376 (50.3%) | 53/379 (14.0%) |
| Total | 616/1295 (47.6%) | 427/1294 (33.0%) | 142/1254 (11.3%) |

**Figure S15.**
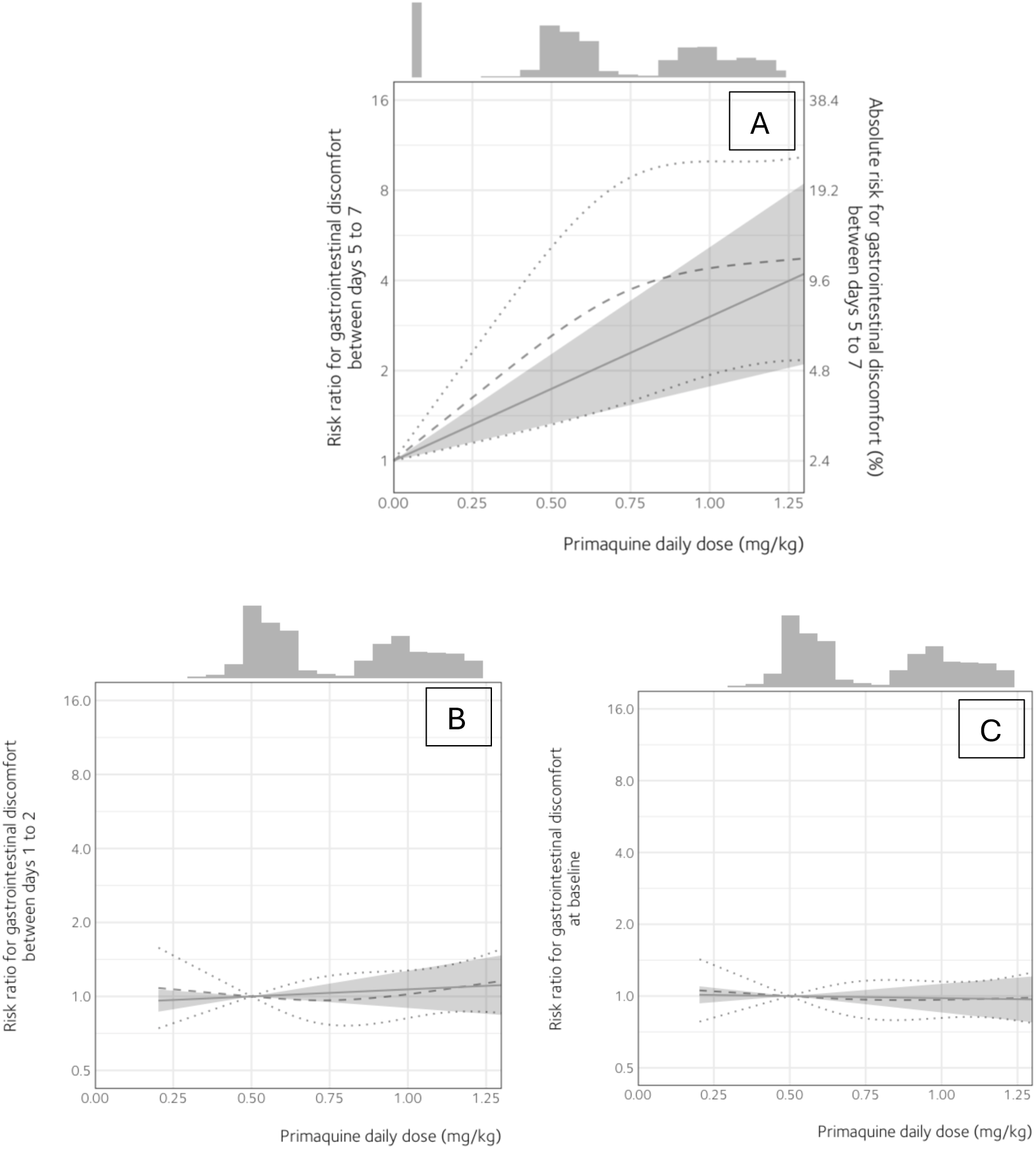
Risk ratio of gastrointestinal discomfort (A) between days 5 to 7, (B) between days 1 to 2, and (C) at day 0 [baseline]. (A) The reference value (RR = 1) was set at the primaquine daily dose of 0 mg/kg. Estimates were derived from a multivariable Poisson model (n = 952). Two different specifications for primaquine daily dose were shown to evaluate potential non-linearity across doses: linear trend (solid grey line with shaded region for 95% CI) and restricted cubic spline with three knots (dashed curve with two dotted curves representing the 95% CI limits). Inclusion of study site (p = 0.7) in a sensitivity analysis did not alter the effect-estimate for dose. The secondary y-axis displays the estimated absolute risk of gastrointestinal discomfort as a percentage. These values are derived by scaling the model-predicted risk ratios by the baseline risk. The histogram along the top margin shows the distribution of primaquine daily doses in the model data, with the leftmost bar representing patients who were treated without primaquine (i.e., 0 mg/kg). The vertical axis is shown on a logarithmic scale. (B, C) The reference value, where the risk ratio equals one, was set at the primaquine daily dose of 0·5 mg/kg. Estimates were derived from a multivariable Poisson model (n in panel A = 767, n in panel B = 767). Two different specifications for primaquine daily dose were shown to evaluate potential non-linearity across doses: trend (solid grey line with shaded region for 95% CI) and restricted cubic spline with three knots (dashed curve with two dotted curves representing the 95% CI limits). The histogram along the top margin shows the distribution of primaquine daily doses in the model data, with the leftmost bar representing patients who were treated without primaquine (i.e., 0 mg/kg). The vertical axis is shown on a logarithmic scale. The y-axis is displayed up to a risk ratio of 16 to enable comparison with the primary gastrointestinal tolerability endpoint.

**Table S9.** Pooled counts and percentages of patients who experienced acute vomiting within 1 hour of taking primaquine between days 0 and 14.

| Primaquine daily dose | Days of observation |
| --- | --- |
|  | 0–14 |
| Low | 0/206 (0%) |
| Intermediate | 15/642 (2·3%) |
| High | 14/380 (3·7%) |
| Total | 29/1228 (2·4%) |

**Figure S16.**
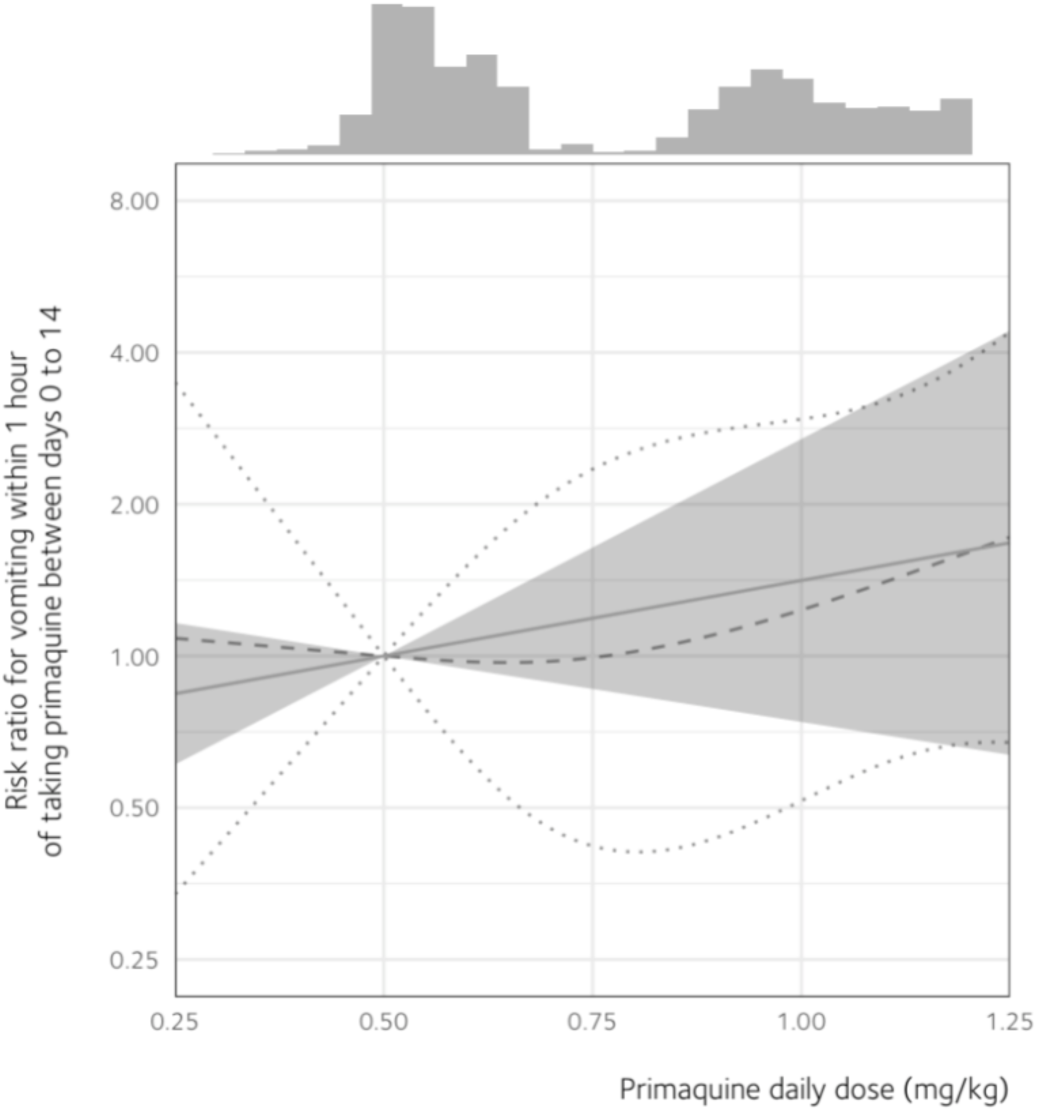
Risk ratio of acute vomiting within 1 hour of taking primaquine between days 0 and 14. The reference value, where the risk ratio equals one, was set at the primaquine daily dose of 0·5 mg/kg. Estimates were derived from a multivariable Poisson model (n = 767). Two different specifications for primaquine daily dose were shown to evaluate potential non-linearity across doses: trend (solid grey line with shaded region for 95% CI) and restricted cubic spline with three knots (dashed curve with two dotted curves representing the 95% CI limits). The histogram along the top margin shows the distribution of primaquine daily doses in the model data. The vertical axis is shown on a logarithmic scale.

**Table S10.** Poisson model estimates with robust standard errors.

| Model | RR (95% robust CI) |
| --- | --- |
| **Tolerability** |  |
| Effect of increasing 0·25 mg/kg daily dose on GI discomfort (days 5–7) | 1·32 (1·17 to 1·48) |
| Effect of increasing 0·25 mg/kg daily dose on GI discomfort (day 0) | 0·99 (0·96 to 1·02) |
| Effect of increasing 0·25 mg/kg daily dose on GI discomfort (days 1–2) | 1·03 (0·97 to 1·10) |
| Effect of increasing 0·25 mg/kg daily dose on acute vomiting | 1·19 (0·84 to 1·67) |
| **Safety** |  |
| Effect of increasing 0·25 mg/kg daily dose on clinical anaemia | 0·90 (0·80 to 1·01) |

**Figure S17.**
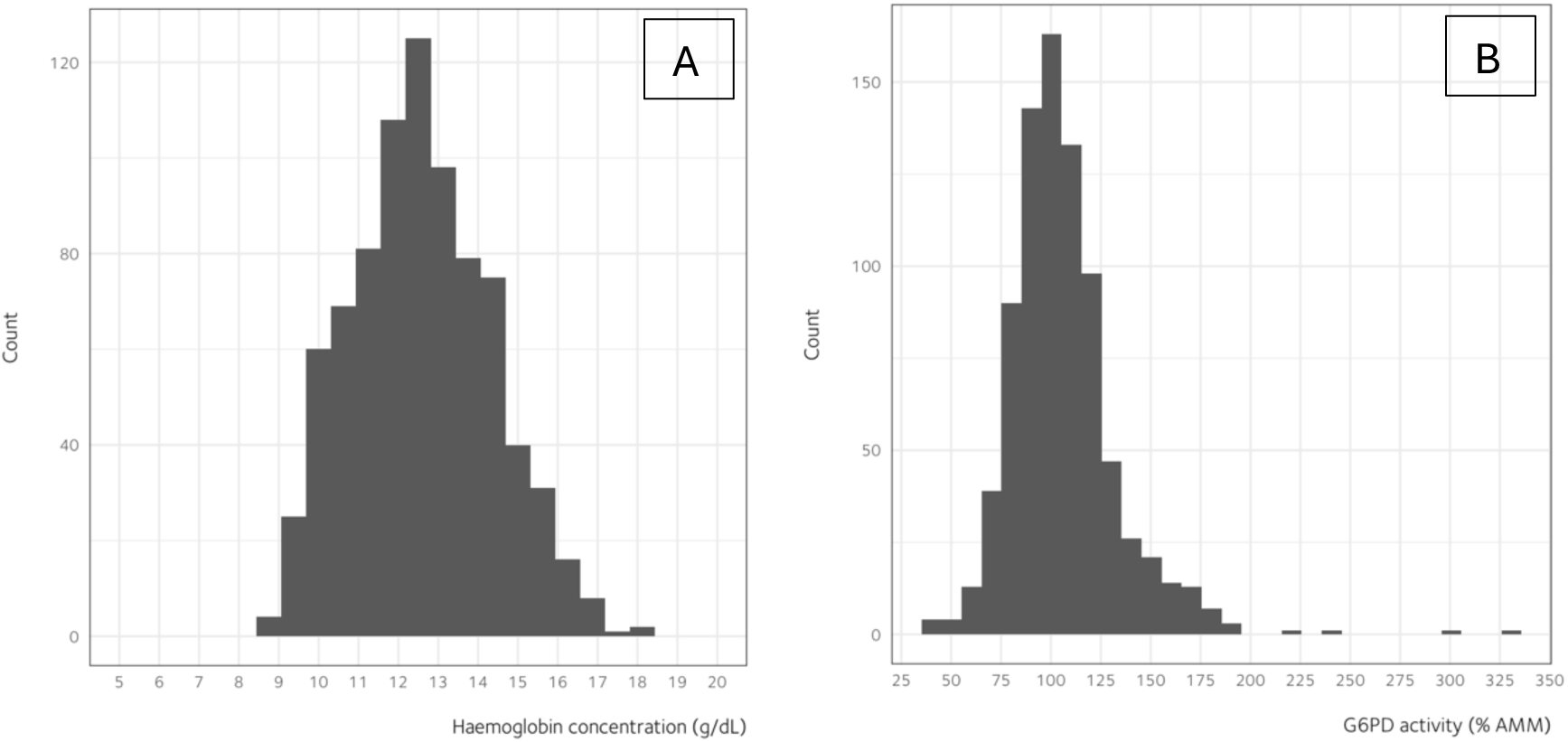
**(A) Haemoglobin concentrations and (B) G6PD activity levels at patient enrolment in the primary safety dataset (n = 822)**

**Table S11.** Haematological adverse events on days 1–14, by dose group.

| Haematological adverse event | Primaquine daily dose |  |  |  |
| --- | --- | --- | --- | --- |
|  | No primaquine | Low | Intermediate | High |
| Haemoglobin fall to <5 g/dL | 0/173 | 0/6 | 0/318 | 0/325 |
| Haemoglobin fall >5 g/dL from baseline | 0/173 | 0/6 | 0/318 | 0/325 |
| Blood transfusion | 0/173 | 0/6 | 0/318 | 0/325 |
| Renal failure requiring dialysis | 0/173 | 0/6 | 0/318 | 0/325 |
| Death | 0/173 | 0/6 | 0/318 | 0/325 |

**Figure S18.**
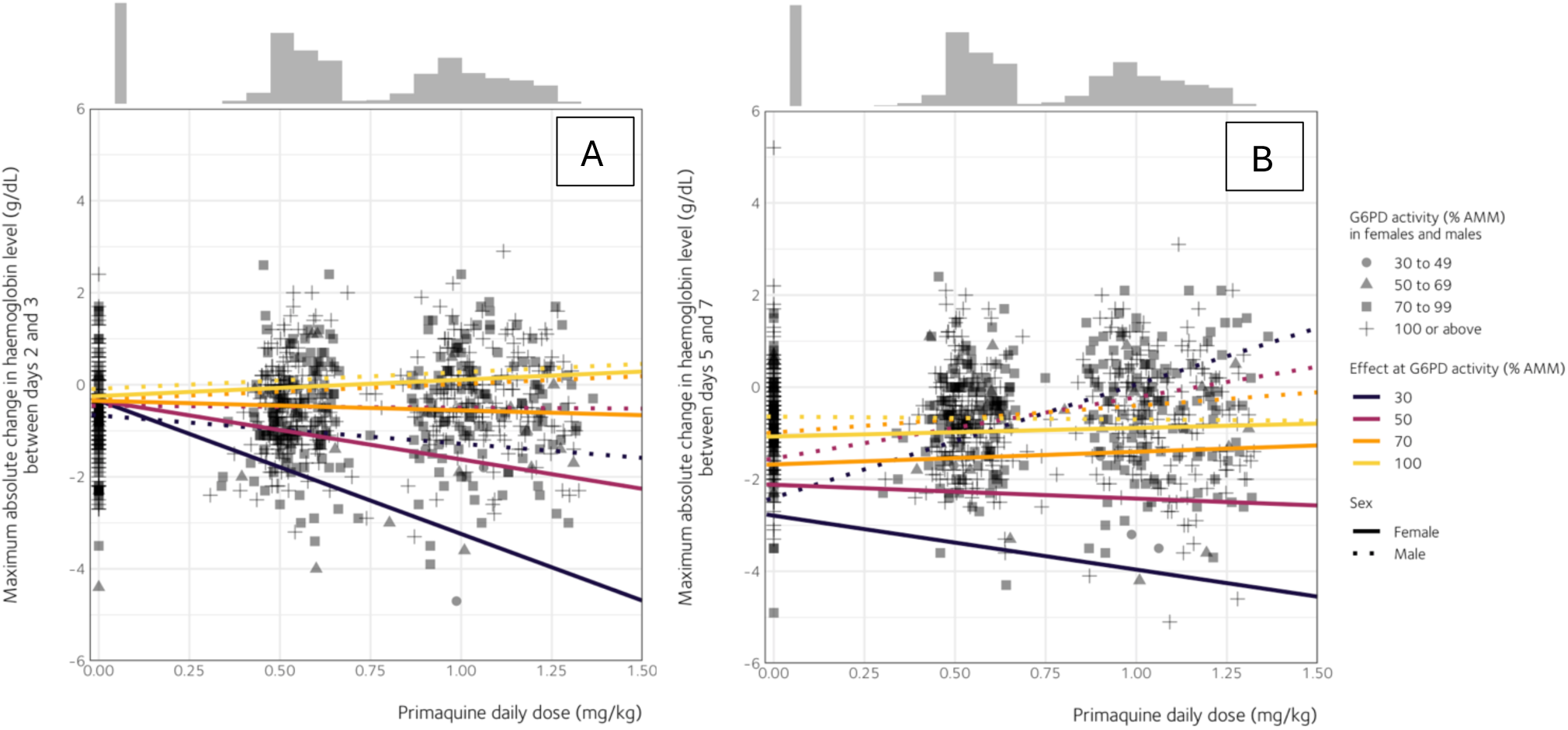
Maximum absolute change from baseline in haemoglobin levels (A) between days 2 and 3 and (B) between days 5 and 7 across primaquine daily doses, sex, and G6PD activity levels. Estimates were derived from a multivariable linear model fitted to the subsets of the primary safety dataset (n in panel A = 794, n in panel B = 781). An interaction term was included in the model to account for differential treatment effects of primaquine daily dose as sex and G6PD activity vary. A restricted cubic spline was specified for G6PD activity to model a non-linear trend. Primaquine daily dose and G6PD activity were both modelled as continuous covariates. The plot shows model-implied predictions only at four selected cut-offs (30%, 50%, 70%, and 100%) of G6PD activity to aid interpretation. The histogram along the top margin shows the distribution of primaquine daily doses in the model data, with the leftmost bar representing patients who were treated without primaquine (i.e., 0 mg/kg). The vertical axis is shown on a logarithmic scale. AMM, adjusted male median.

**Figure S19.**
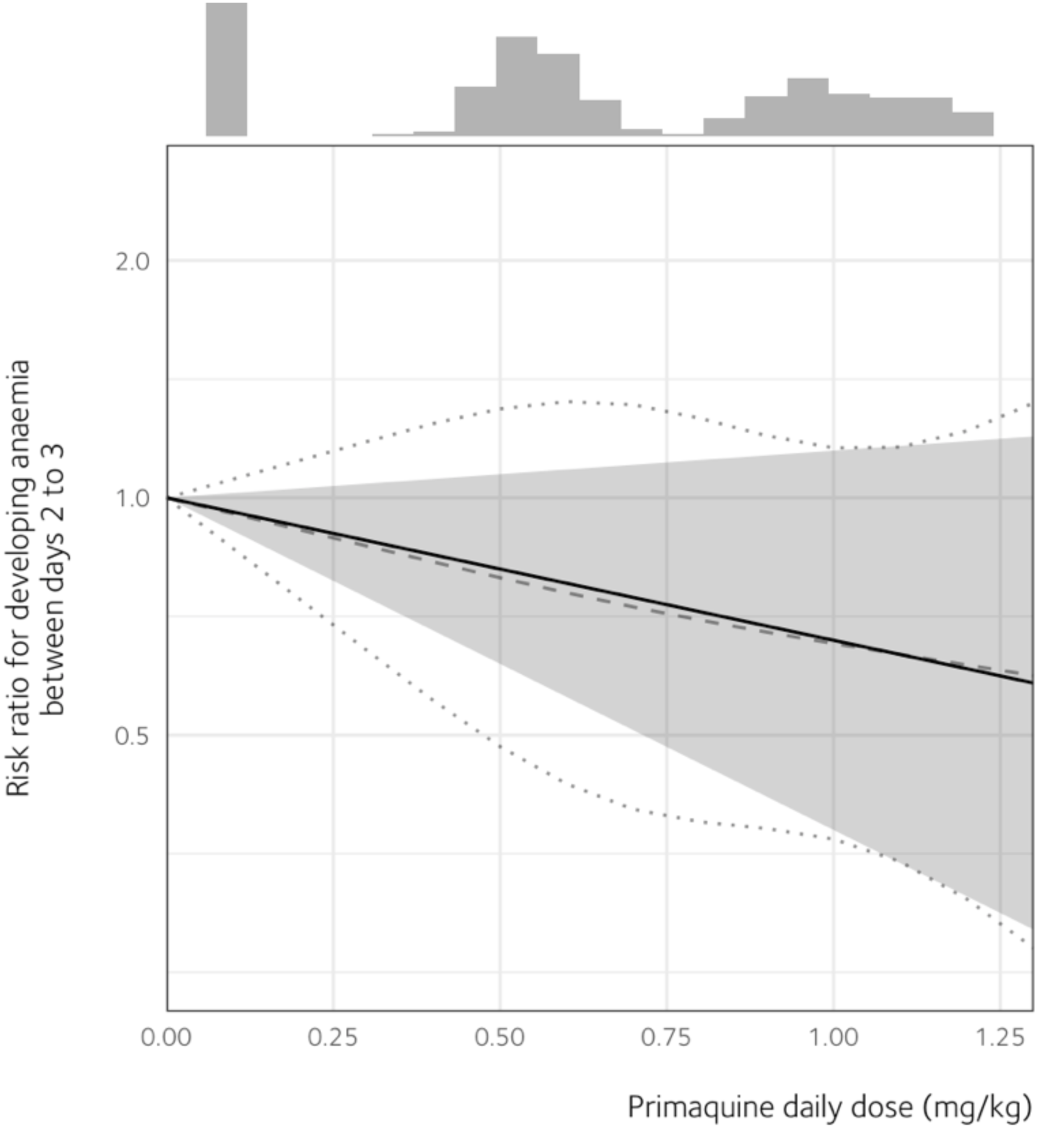
Risk ratio of developing anaemia (haemoglobin <11 g/dL) between days 2 to 3 in patients with G6PD activity of at least 70% and had baseline haemoglobin level of at least 11 g/dL. The reference value, where the risk ratio equals one, was set at the primaquine daily dose of 0 mg/kg. Estimates were derived from a multivariable Poisson model, fitted to a subset of the primary tolerability dataset. Two different specifications for primaquine daily dose were shown to evaluate potential non-linearity across doses: linear trend (solid grey line with shaded region for 95% CI) and restricted cubic spline with three knots (dashed curve with two dotted curves representing the 95% CI limits). The histogram along the top margin shows the distribution of primaquine daily doses in the model data (n = 612), with the leftmost bar representing patients who were treated without primaquine (i.e., 0 mg/kg). The vertical axis is shown on a logarithmic scale.

**Figure S20.**
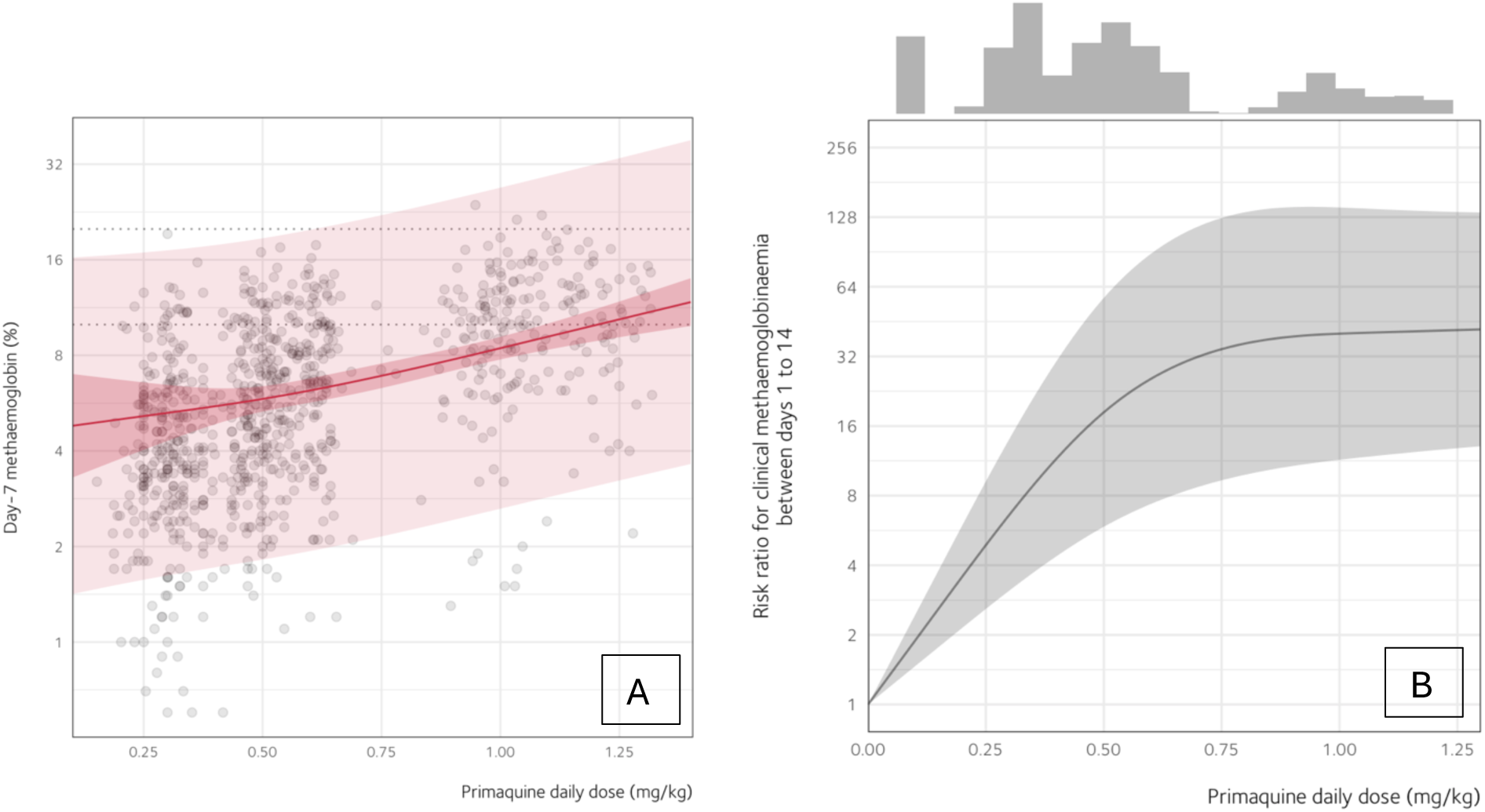
Effect of primaquine daily dose on methaemoglobin levels. (B) Effect of primaquine daily dose on day 7 methaemoglobin levels (n = 886), modelled using multivariable linear regression, adjusting for age, sex, baseline parasite density, and study site. A restricted cubic spline with three knots was applied to primaquine daily dose. The solid curve represents the expected day 7 methaemoglobin levels across the range of primaquine daily doses. The thick and thin shaded regions represent the 95% confidence intervals and prediction intervals, respectively. Each dot represents an individual patient’s day 7 methaemoglobin level. The dotted horizontal lines have y-intercepts of 10% and 20%, thresholds at which symptoms related to tissue hypoxia (e.g., light-headedness, tachycardia) may become more apparent. (B) Risk ratios of clinical methaemoglobinaemia (i.e., 10% or more) from day 1 to 14 across primaquine daily doses. Estimates were modelled using a multivariable Poisson model with a restricted cubic spline with three knots applied to primaquine daily dose. The solid curve and shaded region represent the point estimates and 95% confidence intervals, respectively. The histogram along the top margin shows the distribution of primaquine daily doses in the model data (n = 1026), with the leftmost bar representing patients who were treated without primaquine (i.e., 0 mg/kg). The vertical axis is shown on a logarithmic scale.

## References

1. Price RN, Commons RJ, Battle KE, Thriemer K, Mendis K. *Plasmodium vivax* in the era of the shrinking *P. falciparum* map. Trends Parasitol 2020; 36(6): 560–70.

2. Kementerian Kesehatan Republik Indonesia. Laporan situasi terkini perkembangan program pengendalian malaria di Indonesia tahun 2024, 2025.

3. Djaafara BA, Sherrard-Smith E, Churcher TS, et al. Spatiotemporal heterogeneity in malaria transmission across Indonesia: analysis of routine surveillance data 2010–2019. BMC Med 2025; 23(1): 136.

4. Fadilah I, Djaafara BA, Lestari KD, et al. Quantifying spatial heterogeneity of malaria in the endemic Papua region of Indonesia: Analysis of epidemiological surveillance data. Lancet Reg Health Southeast Asia 2022; 5.

5. Phyo AP, Dahal P, Mayxay M, Ashley EA. Clinical impact of vivax malaria: a collection review. PLoS Med 2022; 19(1): e1003890.

6. Commons RJ, Rajasekhar M, Edler P, et al. Effect of primaquine dose on the risk of recurrence in patients with uncomplicated *Plasmodium vivax*: a systematic review and individual patient data meta-analysis. Lancet Infect Dis 2023.

7. Rajasekhar M, Simpson JA, Ley B, et al. Primaquine dose and the risk of haemolysis in patients with uncomplicated *Plasmodium vivax* malaria: a systematic review and individual patient data meta-analysis. Lancet Infect Dis 2023.

8. World Health Organization. WHO guidelines for malaria, 30 November 2024, 2024.

9. Kementerian Kesehatan Republik Indonesia. Buku saku tatalaksana kasus malaria; 2023.

10. Sadhewa A, Cassidy-Seyoum S, Acharya S, et al. A review of the current status of G6PD deficiency testing to guide radical cure treatment for vivax malaria. Pathogens 2023; 12(5): 650.

11. Nelwan EJ, Ekawati LL, Tjahjono B, et al. Randomized trial of primaquine hypnozoitocidal efficacy when administered with artemisinin-combined blood schizontocides for radical cure of *Plasmodium vivax* in Indonesia. BMC Med 2015; 13: 294.

12. Poespoprodjo JR, Burdam FH, Candrawati F, et al. Supervised versus unsupervised primaquine radical cure for the treatment of falciparum and vivax malaria in Papua, Indonesia: a cluster-randomised, controlled, open-label superiority trial. Lancet Infect Dis 2022; 22(3): 367–76.

13. Sutanto I, Tjahjono B, Basri H, et al. Randomized, Open-Label Trial of Primaquine against Vivax Malaria Relapse in Indonesia. Antimicrob Agents Chemother 2013; 57(3): 1128–35.

14. Pasaribu AP, Chokejindachai W, Sirivichayakul C, et al. A randomized comparison of dihydroartemisinin-piperaquine and artesunate-amodiaquine combined with primaquine for radical treatment of vivax malaria in Sumatera, Indonesia. J Infect Dis 2013; 208(11): 1906–13.

15. Taylor WRJ, Thriemer K, Von Seidlein L, et al. Short-course primaquine for the radical cure of *Plasmodium vivax* malaria: a multicentre, randomised, placebo-controlled non-inferiority trial. Lancet 2019; 394(10202): 929–38.

16. Commons RJ, Thriemer K, Humphreys G, et al. The Vivax Surveyor: Online mapping database for *Plasmodium vivax* clinical trials. Int J Parasitol Drugs Drug Resist 2017; 7(2): 181–90.

17. Infectious Diseases Data Observatory (IDDO). IDDO SDTM implementation manual. 2023. https://www.iddo.org/tools-and-resources/data-tools (accessed: 01/04/2024).

18. Stewart LA, Clarke M, Rovers M, et al. Preferred Reporting Items for Systematic Review and Meta-Analyses of individual participant data: the PRISMA-IPD Statement. JAMA 2015; 313(16): 1657–65.

19. World Health Organization. Technical consultation to review the classification of glucose-6-phosphate dehydrogenase (G6PD), 2022.

20. Zou G. A modified poisson regression approach to prospective studies with binary data. American journal of epidemiology 2004; 159(7): 702–6.

21. Sterne JA, Savović J, Page MJ, et al. RoB 2: a revised tool for assessing risk of bias in randomised trials. BMJ 2019; 366.

22. Sterne JA, Hernán MA, Reeves BC, et al. ROBINS-I: a tool for assessing risk of bias in non-randomised studies of interventions. BMJ 2016; 355.

23. WorldWide Antimalarial Resistance Network (WWARN). Primaquine Indonesia study group. 2023. https://www.iddo.org/primaquine-indonesia-study-group (accessed: 27/02/2025).

24. Hasugian A, Purba H, Kenangalem E, et al. Dihydroartemisinin-piperaquine versus artesunate-amodiaquine: superior efficacy and posttreatment prophylaxis against multidrug-resistant *Plasmodium falciparum* and *Plasmodium vivax* malaria. Clin Infect Dis 2007; 44(8): 1067–74.

25. Lidia K, Dwiprahasto I, Kristin E. Therapeutic Effects of Dyhidroartemisinin Piperaquine Versus Chloroquine for Uncomplicated Vivax Malaria in Kupang, East Nusa Tenggara, Indonesia. Age 2015; 13: 50.

26. Sutanto I, Soebandrio A, Ekawati LL, et al. Tafenoquine co-administered with dihydroartemisinin-piperaquine for the radical cure of *Plasmodium vivax* malaria (INSPECTOR): a randomised, placebo-controlled, efficacy and safety study. Lancet Infect Dis 2023.

27. Maguire JD, Krisin, Marwoto H, Richie TL, Fryauff DJ, Baird JK. Mefloquine is highly efficacious against chloroquine-resistant *Plasmodium vivax* malaria and Plasmodium falciparum malaria in Papua, Indonesia. Clin Infect Dis 2006; 42(8): 1067–72.

28. Arcelia F, Pasaribu AP, Yanni GN. Effectiveness of dihydroartemisinin-piperaquine after 10 years as treatment for vivax malaria in Indonesia. J Infect Dev Ctries 2023; 17(05): 700–6.

29. Chu CS, Bancone G, Moore KA, et al. Haemolysis in G6PD heterozygous females treated with primaquine for *Plasmodium vivax* malaria: a nested cohort in a trial of radical curative regimens. PLoS Med 2017; 14(2): e1002224.

30. Lipsitch M, Tchetgen ET, Cohen T. Negative controls: a tool for detecting confounding and bias in observational studies. Epidemiol 2010; 21(3): 383–8.

31. Rajgor D, Gogtay N, Kadam V, et al. Antirelapse efficacy of various primaquine regimens for *Plasmodium vivax*. Malar Res Treat 2014; 2014(1): 347018.

32. Saravu K, Tellapragada C, Kulavalli S, et al. A pilot randomized controlled trial to compare the effectiveness of two 14-day primaquine regimens for the radical cure of vivax malaria in South India. Malar J 2018; 17: 1–11.

33. Chamma-Siqueira NN, Negreiros SC, Ballard S-B, et al. Higher-dose primaquine to prevent relapse of *Plasmodium vivax* malaria. N Engl J Med 2022; 386(13): 1244–53.

34. Eng V, Lek D, Sin S, et al. 14 days of high-dose versus low-dose primaquine treatment in patients with *Plasmodium vivax* infection in Cambodia: a randomised, single-centre, open-label efficacy study. Lancet Infect Dis 2025.

35. Degaga TS, Pasaribu AP, Tripura R, et al. Effectiveness and safety of high dose primaquine and tafenoquine in Plasmodium vivax patients (EFFORT)-a multi-centre, open label, superiority randomised controlled trial. Lancet Infect Dis 2026.

36. Collins WE, Jeffery GM. Primaquine resistance in *Plasmodium vivax*. Am J Trop Med Hyg 1996; 55(3): 243–9.

37. Garrison PL, Hankey DD, Coker WG, et al. 2. CURE OF KOREAN VIVAX MALARIA WITH PAMAQUINE AND PRIMAQUINE. J Am Med Assoc 1952; 149(17): 1562–3.

38. White NJ. Determinants of relapse periodicity in *Plasmodium vivax* malaria. Malar J 2011; 10(1): 297.

39. Lestari KD, Surendra H, Djaafara BA, et al. Epidemiology of malaria and district-level factors associated with malaria elimination in Sumatra region, Indonesia: a retrospective analysis of surveillance data. medRxiv 2025: 2025.08. 28.25333185.

40. Mehdipour P, Rajasekhar M, Dini S, et al. Effect of adherence to primaquine on the risk of *Plasmodium vivax* recurrence: a WorldWide Antimalarial Resistance Network systematic review and individual patient data meta-analysis. Clin Infect Dis 2023; 22(1): 306.

41. Fryauff D, Baird K, Basri H, et al. Randomised placebo-controlled trial of primaquine for prophylaxis of falciparum and vivax malaria. Lancet 1995; 346(8984): 1190–3.

42. Clayman CB, Arnold J, Hockwald RS, Yount EH, Edgcomb JH, Alving AS. 3. Toxicity of primaquine in caucasians. J Am Med Assoc 1952; 149(17): 1563–8.

43. Baird JK, Hoffman SL. Primaquine therapy for malaria. Clin Infect Dis 2004; 39(9): 1336–45.

44. Fadilah I, Commons RJ, Chau NH, et al. Methaemoglobin as a surrogate marker of primaquine antihypnozoite activity in *Plasmodium vivax* malaria: A systematic review and individual patient data meta-analysis. PLoS Med 2024; 21(9): e1004411.

45. White NJ, Watson JA, Baird JK. Methaemoglobinaemia and the radical curative efficacy of 8-aminoquinoline antimalarials. Br J Clin Pharmacol 2022; 88(6): 2657–64.

46. Watson JA, Commons RJ, Tarning J, et al. The clinical pharmacology of tafenoquine in the radical cure of *Plasmodium vivax* malaria: An individual patient data meta-analysis. Elife 2022; 11.

47. Green JA, Mohamed K, Goyal N, et al. Pharmacokinetic interactions between tafenoquine and dihydroartemisinin-piperaquine or artemether-lumefantrine in healthy adult subjects. Antimicrob Agents Chemother 2016; 60(12): 7321–32.

48. Commons RJ, Simpson JA, Watson J, White NJ, Price RN. Estimating the Proportion of *Plasmodium vivax* Recurrences Caused by Relapse: A Systematic Review and Meta-Analysis. Am J Trop Med Hyg 2020; 103(3): 1094–9.

49. Battle KE, Lucas TCD, Nguyen M, et al. Mapping the global endemicity and clinical burden of *Plasmodium vivax*, 2000–17: a spatial and temporal modelling study. Lancet 2019; 394(10195): 332–43.

50. Becker RAW, A. R.; Brownrigg, R.; Minka, T. P.; Deckmyn, A. maps: Draw Geographical Maps. 3.4.1 ed: R package; 2023.

51. Wickham HC, W.; Henry, L.; Pedersen, T. L.; Takahashi, K.; Wilke, C.; Woo, K.; Yutani, H. ggplot2: Create Elegant Data Visualisations Using the Grammar of Graphics. 3.5.1 ed: R package; 2023.

52. Battle KE, Karhunen MS, Bhatt S, et al. Geographical variation in *Plasmodium vivax* relapse. Malar J 2014; 13(1): 144.

